# Microbial traits have minimal power for predicting death from bloodstream infection

**DOI:** 10.64898/2026.10.08.26365081

**Authors:** Rory Gilliland, Mario E. Valdés-Tresanco, Andriy Plakhotnyk, Anika Westlund, Joshua T. Smith, Thi Mui Pham, Tatum D. Mortimer, Bruce J. Walker, Ashlee M. Earl, Yonatan H. Grad, M. Ethan MacDonald, Ian A. Lewis, the PRIME Consortium

## Abstract

Bloodstream infection (BSI) is a life-threatening condition where the risk of dying is affected by both patient-and pathogen-related factors. Certain microbial traits (e.g., virulence genes) are thought to affect survival rates from BSIs, but the magnitude of their impact on human mortality remains unclear. To address this, we analyzed the microbial genomes, quantitative proteomes, and electronic medical records (EMRs) from 35,746 BSIs from Calgary, Canada (2006–2022). We show that EMR-based machine learning models accurately predict 30-day mortality (AUC=0.83), but surprisingly, integrating >70,000 microbial genes/proteins does not improve this performance. We demonstrate that microbial factors co-segregate with specific patient cohorts and are therefore not independent predictors of risk. We conclude that microbial virulence factors have minimal impact on mortality rates from BSIs.

**One-Sentence Summary:** Microbial genes and protein abundance levels have minimal power for predicting 30-day mortality in bloodstream infections.

## Main Text

There are more than 500,000 bloodstream infections (BSIs) each year in North America resulting in over 90,000 deaths (*1*). The specific risks each BSI patient faces are affected by a wide range of patient-and pathogen-related factors. Patient-related factors such as age (*2–4*), immune function (*5–8*), comorbidities (e.g., cancer, heart disease, diabetes, etc.) (*2*, *4*, *9*), and acute clinical presentation (e.g., blood pressure, respiratory rate, etc.) (*4*, *9*, *10*) have each been linked to differential survival rates. Pathogen-related factors such as the species (*11*), strain (*12*, *13*), antimicrobial resistance phenotype (*4*, *14*), and the presence of microbial virulence factors (*4*, *15–23*) are also thought to modulate survival rates in BSIs. These risk factors are non-trivial. For example, the species of pathogen has been linked to a ∼30% difference in 30-day mortality in BSIs (*11*), and BSI mortality risk has been shown to increase by 2% with each year of patient age (*2*). Similarly, the presence of hypervirulence traits (e.g., *purR* mutants in *Staphylococcus aureus*) can increase 1-week mortality by ∼75% in animal models (*24*). Although hundreds of individual microbial risk factors like these have now been identified, a central question remains unanswered: to what extent do these microbial factors contribute to a patient’s risk of dying from a BSI? The goal of this study was to answer this question.

Recent advances in machine learning have leveraged electronic medical records (EMRs) to build accurate models of sepsis occurrence (AUCs=0.90–0.95) (*25–35*) and BSI outcome (AUCs=0.80–0.85) (*36–42*). Though effective, these models appear to be approaching the limits of predictive performance achievable with EMR data sources alone. Recent studies of human serum from sepsis patients have suggested that the human response to infection may provide additional predictive information (*5*, *43*). However, no equivalent study has been completed to assess the impact of microbial factors on mortality risk. Given the significant associations between microbial traits and BSI outcome, we hypothesized that an integrated patient/microbe model would provide superior performance and facilitate new insights into the relationship between host and microbial factors in shaping the clinical trajectory of infections.

## Results

### Calgary BSI Cohort

Forecasting patient outcomes for BSIs is complicated by the extensive list of mortality-linked traits, the co-segregation of risk factors, and the limited availability of medical records that are linked to molecular data of pathogens. To overcome this challenge, we collected the *Calgary BSI Cohort*, a dataset of 35,746 (Fig. S1) isolates representing every BSI observed in the Calgary Health Region (Canada) between 2006 and 2022. To assess the impact of microbial traits on outcome, we analyzed the multi-omics profile of every *Escherichia coli* (*n*=10,501), *Staphylococcus aureus* (*n*=8,078), *Enterococcus faecium* (*n*=844), and *Klebsiella pneumoniae* (*n*=2,427) isolate from the cohort. We chose to focus on these four pathogens to capture the more pervasive (i.e., *E. coli*, *K. pneumoniae*, *S. aureus*) and higher risk (i.e., *E. faecium*) causative agents of BSIs. These analyses included whole genome sequencing (Illumina) and tandem mass tag (TMT) liquid chromatography mass spectrometry (LC-MS) proteomics analyses of each isolate. Pan genomes and pan proteomes (inclusive of all core and accessory genes present in at least 1% of isolates) were then compiled for each species and the presence/absence of genes was recorded, which resulted in 8,863–68,192 genomic features (depending on the species). In addition, 1,805–2,672 protein abundance levels (depending on the species) were captured per organism. Molecular data from each isolate was matched to the 3,962 EMR fields including patient age, sex, Charlson Comorbidity Indices (CCI), Pitt Bacteremia Scores (PBS), Sequential Organ Failure Assessment (SOFA) scores, International Classification of Disease, Tenth Revision, Canadian version (ICD-10-CA) codes, and laboratory data such as blood urea nitrogen (Table S1).

### Experimental Strategy

To better understand the clinical impact of both patient and microbial factors in BSIs, we created machine learning (ML) models that included either microbial features, EMR features, or combinations of both (Fig. S2). These models were generated using a grid search-and cross-validation-based model selection process that considered over one million combinations of a diverse array of preprocessing techniques, feature selection methods, classification algorithms, and hyperparameters thereof. The performance of each model was evaluated using a broad suite of metrics including AUC, sensitivity, specificity, and F1 score. We identified the most important contributors to these models using independent held-out performance estimation and model explainability techniques (i.e. Shapley additive explanations, SHAP) (*44*, *45*). This approach facilitated detailed quantitative insights into the contributions of each factor to predictions of 30-day mortality.

### Patient-Related Predictors of Mortality

The role that patient-related factors play in shaping the severity of BSIs has been studied intensively (*2–4*, *9*, *10*, *36–42*). These studies have shown that age, comorbidities, and acute clinical factors are significantly linked to 30-day mortality and can be used to create accurate predictive ML models of outcome. Our epidemiological records suggest that these relationships are similar in the Calgary region (Fig. 1). For example, patient age is directly proportional to 30-day mortality in BSI patients (Fig. 2a). However, since regional demographic differences can affect BSI epidemiology (*1*, *3*), we constructed a patient-only ML model using the *Calgary BSI Cohort* data to identify the primary host contributions to 30-day mortality in the study population. These predictive models were then used as a baseline to assess host-versus-microbial contributions to outcome in our subsequent studies. Our patient-only models were built from an initial set of 3,962 EMR fields from each patient. We then identified the 99 features that were most predictive of 30-day mortality via ReliefF (*46*). These predictive features were then used to build an XGBoost-based ML model of BSI outcome.

**Fig. 1:**
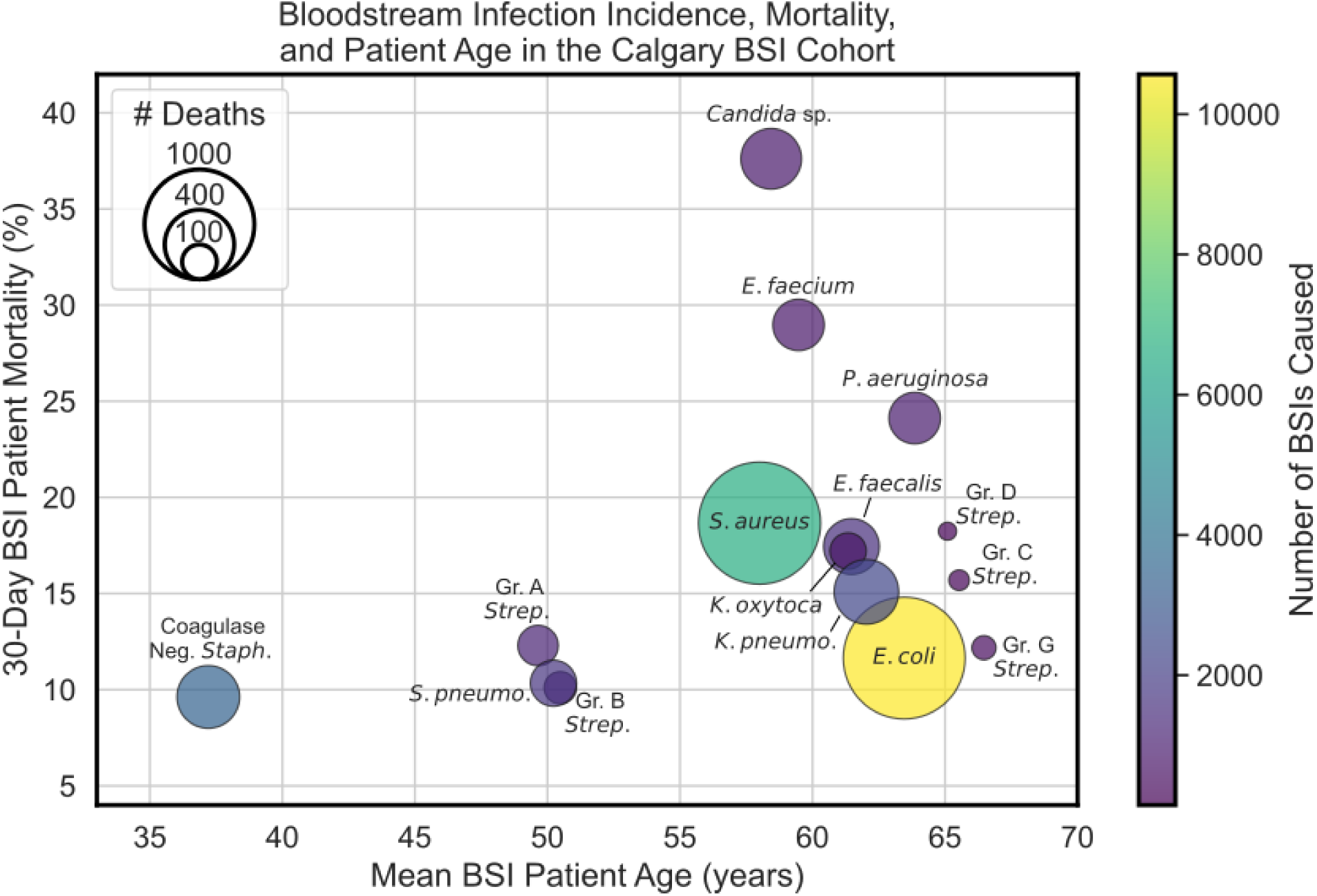
Bloodstream infection incidence, mortality, and patient age by infecting pathogen in the *Calgary BSI Cohort*, a collection of over 35,746 BSI isolates from Calgary, Canada between 2006 and 2022. Inspired by Figure 3 from Verway et al. (*11*). Abbreviations: *E. faecium, Enterococcus faecium; P. aeruginosa, Pseudomonas aeruginosa; K. oxytoca, Klebsiella oxytoca;* Gr. A (-B,-C,-D,-G) *Strep.,* group A (-B,-C,-D,-G) *Streptococcus; E. faecalis, Enterococcus faecalis; K. pneumo, Klebsiella pneumoniae; S. pneumo., Streptococcus pneumoniae;* Coagulase Neg. *Staph,* coagulase negative *Staphylococcus; S. aureus, Staphylococcus aureus; E. coli, Escherichia coli*.

**Fig. 2:**
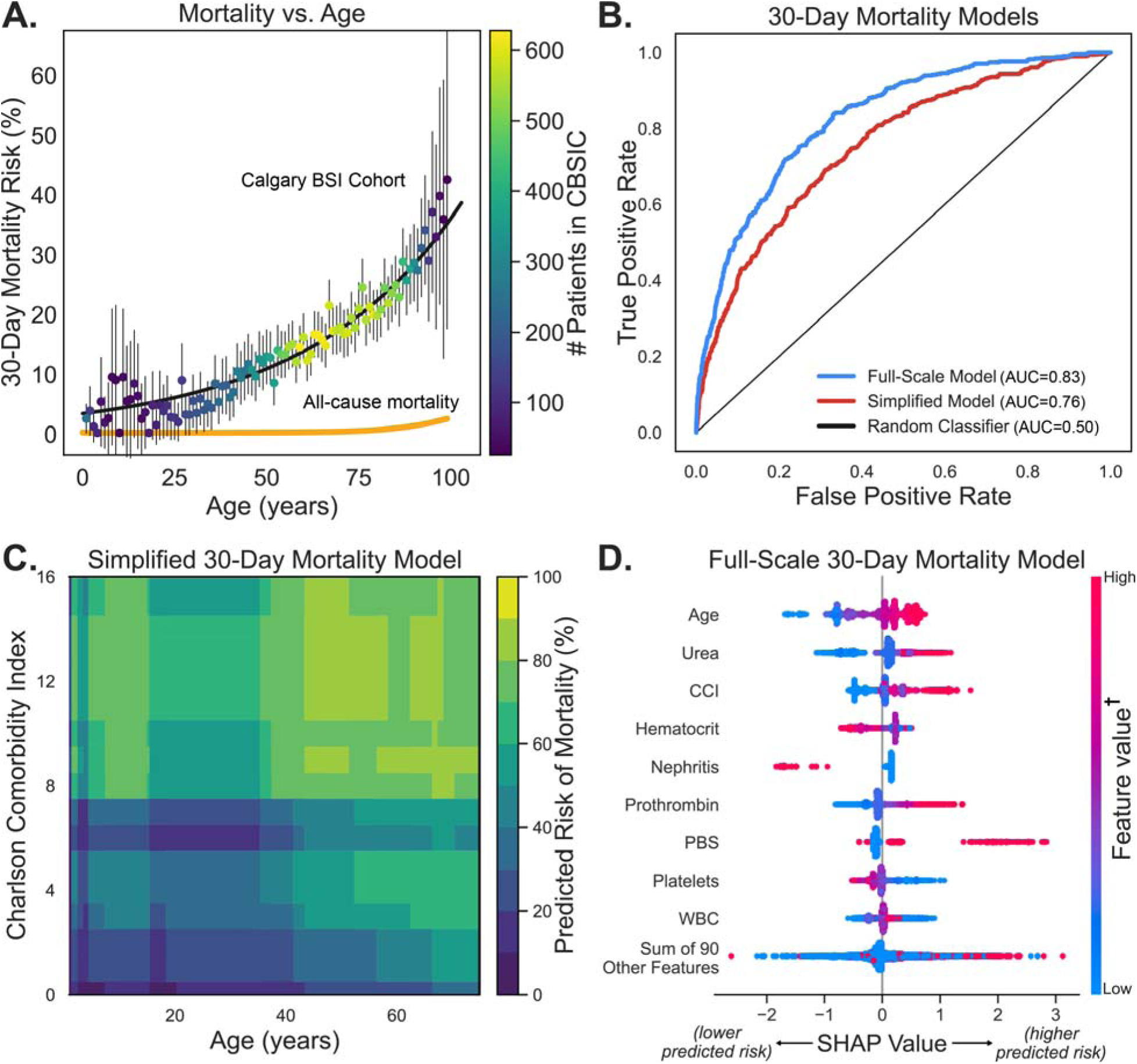
Impact of patient-related factors, extracted from health records, on BSI mortality predictions. **(A)** Relationship between age and the probability of 30-day patient mortality observed in the *Calgary BSI Cohort*, a collection of over 35,746 BSIs from Calgary, Canada between 2006 and 2022. BSI-linked mortality rates (black line) are shown in comparison to average all-cause mortality rates (yellow line) reported in the United States (*79*). **(B)** Receiver operating characteristic curves for 30-day bloodstream infection (BSI) patient mortality models using 99 patient-related features (Full-Scale Model; blue) versus four patient-related features (Simplified Model; red). **(C)** Risk of 30-day BSI patient mortality across a range of patient age and CCI values. PBS and SOFA were fixed to a constant value to enable this visualization. **(D)** SHAP analysis on the full-scale 30-day BSI patient mortality model. Each dot represents a sample in the held-out testing set and is colored by the value of the corresponding feature in that sample. SHAP values are assigned according to their impact on the model’s prediction, where positive SHAP values denote a sample for which the corresponding feature value was interpreted to increase the risk of an adverse clinical outcome by the model, and vice versa. Feature value (†) corresponds to a high value (red) for a given feature (i.e., old age) or low value (blue) for that same feature (i.e., young age). Definitions of patient-related features are provided in Table S1. Abbreviations: AUC, area under the curve; WBC, white blood cell count; Nephritis is an aggregate feature that includes any ICD-10CA code containing the search term “nephritis”.

Our ML model built from EMR records alone mirrored the findings reported by others. On held-out testing data, our model achieved an AUC, balanced accuracy, and F1 score of 0.83, 75%, and 0.438, respectively (Table S2 and Figs. 2b, S3, and S4). These performance metrics match those seen in other large cohorts (AUC 0.80–0.85) (*36–40*, *42*). The most influential features in this model (Fig. 2c-d) included patient age, blood urea nitrogen, CCI (*47*), and PBS (*48*, *49*); each of these features have been reported previously as predictors of BSI outcome (*36–40*, *42*). In addition, we identified “nephritis” (an aggregated feature reflecting any ICD-10 code that includes the keyword’nephritis’; Table S1) as a major predictor of outcome (Fig. 2d). We also constructed models predicting length of hospital stay and healthcare costs linked to each infection using the same feature selection and ML modeling workflow. These models showed that a similar constellation of risk factors contribute to these different dimensions of infection severity (Table S 2 and Fig. S3, S4).

In addition to building predictive models using comprehensive patient records, we also constructed a simplified model using patient-related features that would be available to clinicians in most critical care settings. This compact model only considered patient age, CCI, PBS, and sequential organ failure assessment (SOFA) (*50*). This streamlined model performed only slightly worse than its full-scale counterpart (AUC=0.76; Figs. 2b and c and Table S2).

### Microbial Predictors of Mortality

Our ML models, and those of several groups around the world (*2–4*, *9*, *10*, *36–42*), show that BSI mortality can be predicted from patient factors. However, the performance of these models appears to be limited to an AUC of ∼0.85, suggesting there are significant additional factors shaping the severity of infection. One obvious dimension to BSI outcome that our EMR modeling efforts do not capture are microbial factors, such as virulence and antimicrobial resistance. To address this gap, we developed a set of models that incorporate up to 70,775 microbial features (species classification, sequence type and/or clonal complex, gene presence/absence including the presence of any known resistance or virulence-related genes, and quantitative abundance of proteins in each isolate).

### Importance of Microbial Species in Predicting Mortality

BSIs are caused by a wide range of microbial pathogens. Infections from these diverse species have distinct clinical characteristics with species-specific 30-day mortality rates ranging from 10–37% according to both our data (Fig. 1) and the literature (*11*, *51–53*). Given these well documented species-linked mortality rates (*11*, *51–53*), we anticipated that the performance of our ML models would be significantly improved by including the species of microbe as a predictive feature. To test this, we built ML models of 30-day mortality using patient factors, microbial species, and a combination of both sets of features.

Surprisingly, microbial species information had minimal impact on the predictive performance of our ML models, either alone or when integrated with patient-related factors. When considered alone, models of 30-day mortality using microbial species had an AUC=0.59 – a performance that is only marginally better than a random classifier (*54*). Similarly, when microbial species were added to the patient factors, we observed no change in the predictive power of the models (AUC=0.83 in both cases; Fig. 3a and Table S3). SHAP analysis of the combined patient/microbe models corroborates this finding: although the species information was present in the model, the variance in SHAP values linked to the species was minimal relative to the patient-related predictors (Fig. 3a and b). To more tightly control for patient-related risk factors, we also built models for sub-cohorts constructed from age-, CCI-, and PBS-matched patients. ML models built on these matched cohorts also showed minimal contributions from microbial species to the model performance (Table S3). Collectively, these findings show that microbial species adds little predictive power to models of patient outcome.

**Fig. 3:**
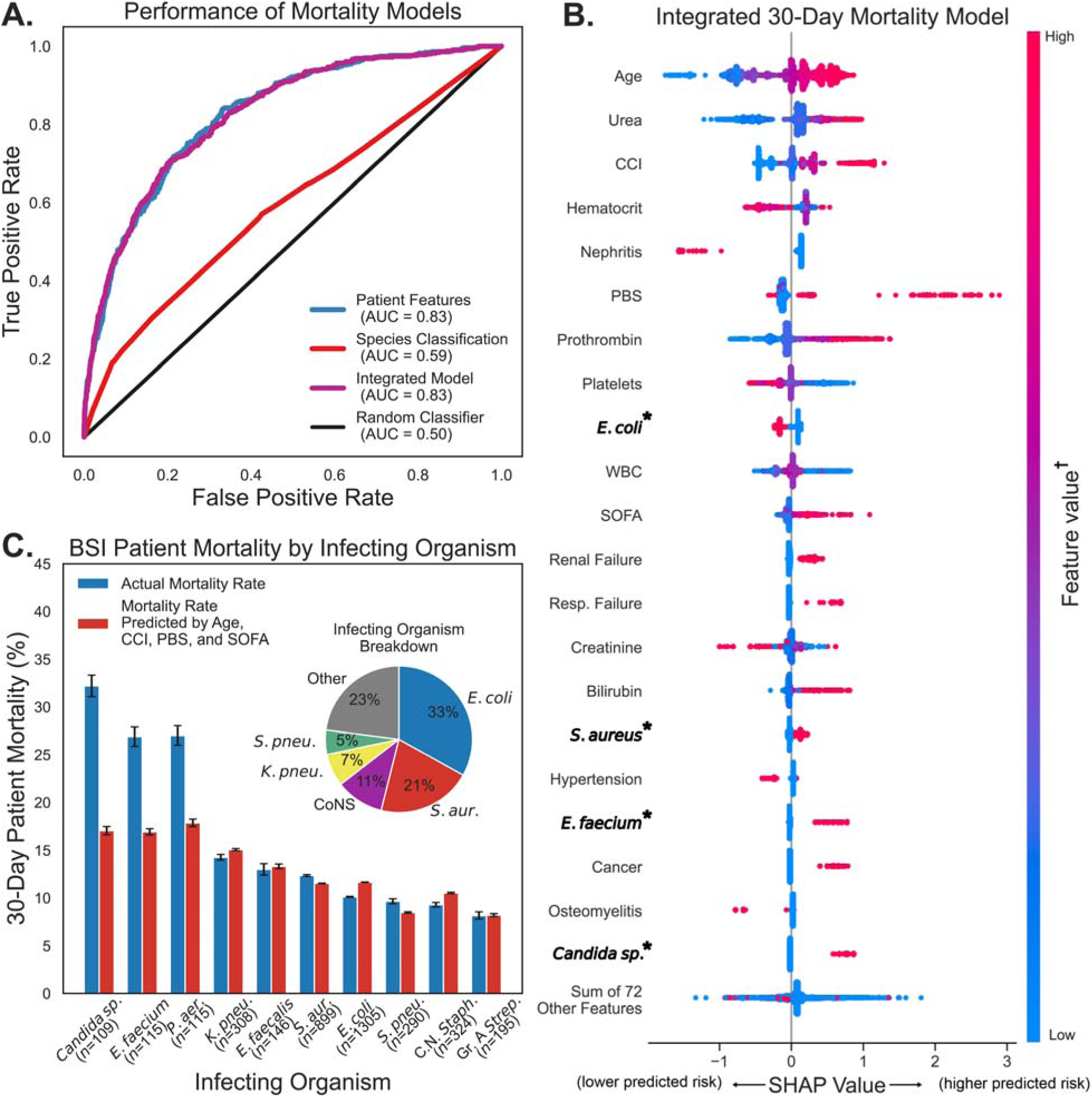
Contribution of patient factors versus BSI pathogen in predicting 30-day mortality. **(A)** Receiver operating characteristic curves for 30-day BSI patient mortality models that include patient-related features (blue), species classification as a feature (red), or a combination of both (purple). **(B)** SHAP analysis on the Integrated Mortality Model. See the caption of Figure 2c for guidance on interpreting this plot. Note that features corresponding to an organism name (e.g., *E. coli*) are encoded as 1 (red dot) if the BSI was caused by that organism. **(C)** Comparison of observed (blue) and predicted (red) 30-day BSI patient mortality rates by infecting organism in a held-out testing dataset. Predicted mortality rates were generated by the Simplified Mortality Model (see Fig. 2b and c), which used patient age, CCI, PBS, and SOFA as features. Note that class weights in the Simplified Mortality Model were not adjusted to generate these predicted mortality rates. Infecting organisms represented by fewer than 100 samples in the testing dataset were excluded from this visualization. Error bars indicate 99% confidence intervals from a 1,000-iteration bootstrap. The inset pie chart depicts a breakdown of BSI incidence by infecting organism. Abbreviations: *S. aur., Staphylococcus aureus; E. faecalis, Enterococcus faecalis;* C. N. *Staph.*, coagulase negative *Staphylococcus; K. pneu., Klebsiella pneumoniae; S. pneu., Streptococcus pneumoniae; P. aer., Pseudomonas aeruginosa;* Gr. A *Strep.,* group A *Streptococcus;* AUC, area under the curve; WBC, white blood cell count.

Given the substantial differences in mortality rates that have been linked to various BSI pathogens both in our data (Fig. 1) and elsewhere (*11*, *51–53*), we speculated that some species-dependent differences in mortality may be attributed to characteristics of the patient population. To test this, we applied our simplified four-factor patient mortality model (detailed above) to cohorts of patients segregated according to their BSI pathogen. These species-segregated predictions of 30-day mortality rates closely matched the *observed* rates (Fig. 3c). Three exceptions to this trend were patients infected with *Candida*, *E. faecium,* or *Pseudomonas aeruginosa—*these groups of patients had higher-than-predicted mortality rates (Fig. 3c). However, for the remaining seven BSI pathogens, our simplified model with just four patient-related factors was sufficient to predict 30-day mortality rates to within ∼2% of their actual values. This finding again suggests that characteristics of the patient are the primary drivers of BSI mortality.

### Importance of Microbial Multi-Omics in Predicting Mortality

An extensive body of literature links the presence of microbial virulence factors and resistance traits to severe infections and higher mortality (*4*, *15–19*, *21*, *22*, *55–58*). Univariate statistical analyses of our data support these observations and identified more than 50 genes and 100 proteins that were significantly linked to differential 30-day mortality (Figs. S5 and S6). One example of these outcome-linked genes is *papG* in *E. coli* (adjusted *p*<0.001 via Chi square test; Fig. 4a-b), a known risk factor for the progression of urinary tract infections into urosepsis (*18*, *21*, *59*). Similarly, the relative abundance of proteins encoded by antimicrobial resistance genes (TetA, PBP2a, ErmA, AphA, and VanX; adjusted *p*<0.05 for all via Mann-Whitney *U* test; Fig. S6) expressed during *in vitro* culture was also linked to higher 30-day mortality in univariate analyses, as has been reported previously (*60–64*). We also identified proteins linked to higher mortality that have not been reported previously. For example, the abundance of *S. aureus* KdpC1, a potassium transporter (*65*), was linked to significantly higher mortality rates (adjusted *p*<0.05 via Mann-Whitney *U* test; Fig. 5a-b).

**Fig. 4:**
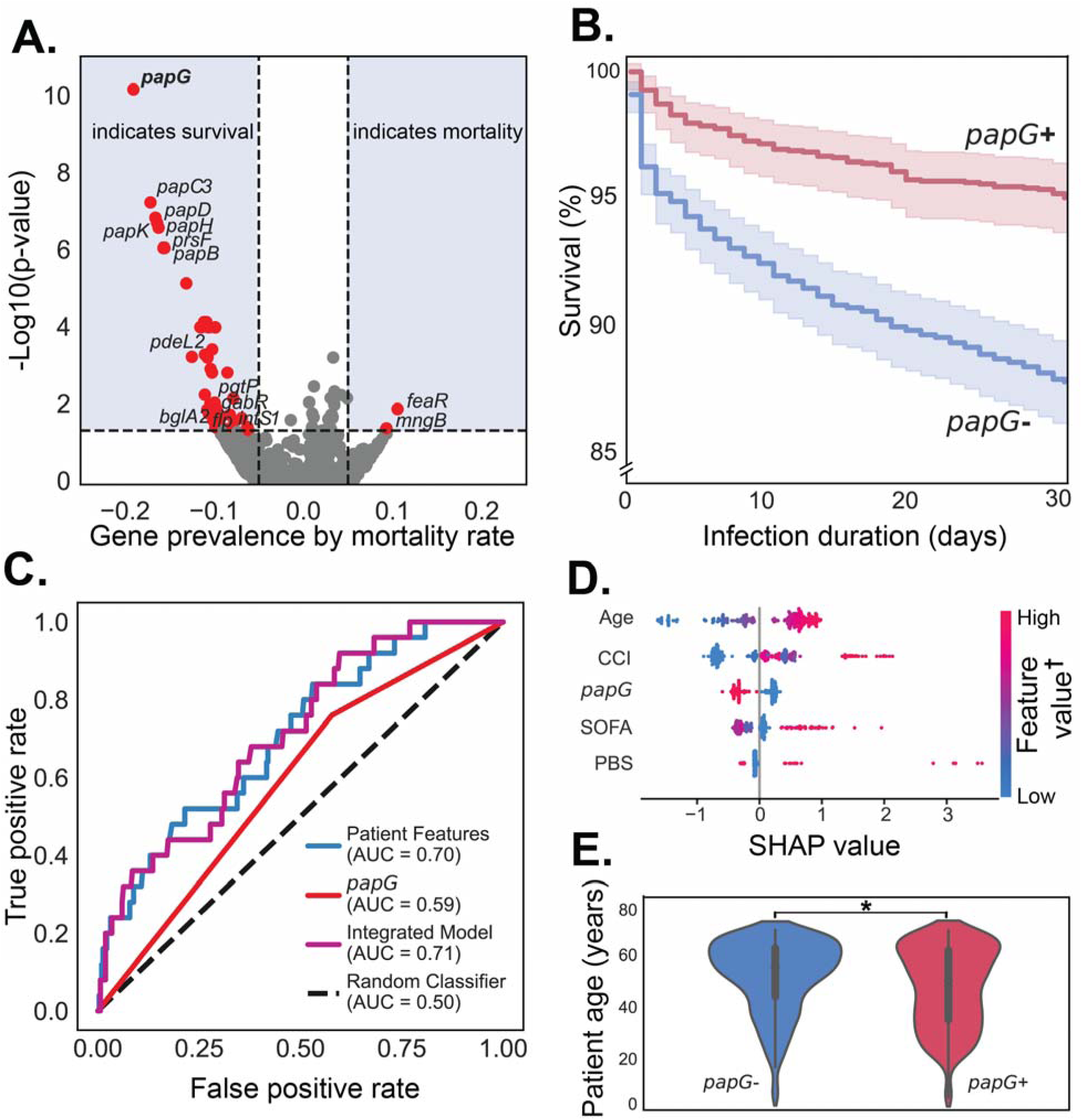
Effect of *E. coli* genes versus patient factors on 30-day BSI mortality predictions. **(A)** Associations between gene presence and 30-day BSI patient mortality in *E. coli* BSIs, with significance determined via chi-square test. Red points in the blue shaded region indicate genes which are significantly associated with mortality (corrected *p*<0.05 after Benjamini-Hochberg; horizontal dashed lines) and have an absolute difference in gene presence proportion exceeding 0.05 (vertical dashed lines). Some red points are not labelled for clarity or due to poor annotation. **(B)** Percent survival from *E. coli* BSIs with *papG* gene presence or absence. **(C)** Receiver operating characteristic curves for models designed to predict 30-day BSI patient mortality using a set of four patient features (age, CCI, PBS, and SOFA), *papG* gene presence, or both. **(D)** SHAP analysis on the integrated patient-and-microbe BSI patient mortality model. See the caption of Figure 2 for guidance on interpreting this plot. **(E)** Relationship between *papG* presence and patient age. Age distributions were compared using a Mann-Whitney *U* test, significance (*p*<0.001) is indicated by an asterisk (*).

**Fig. 5:**
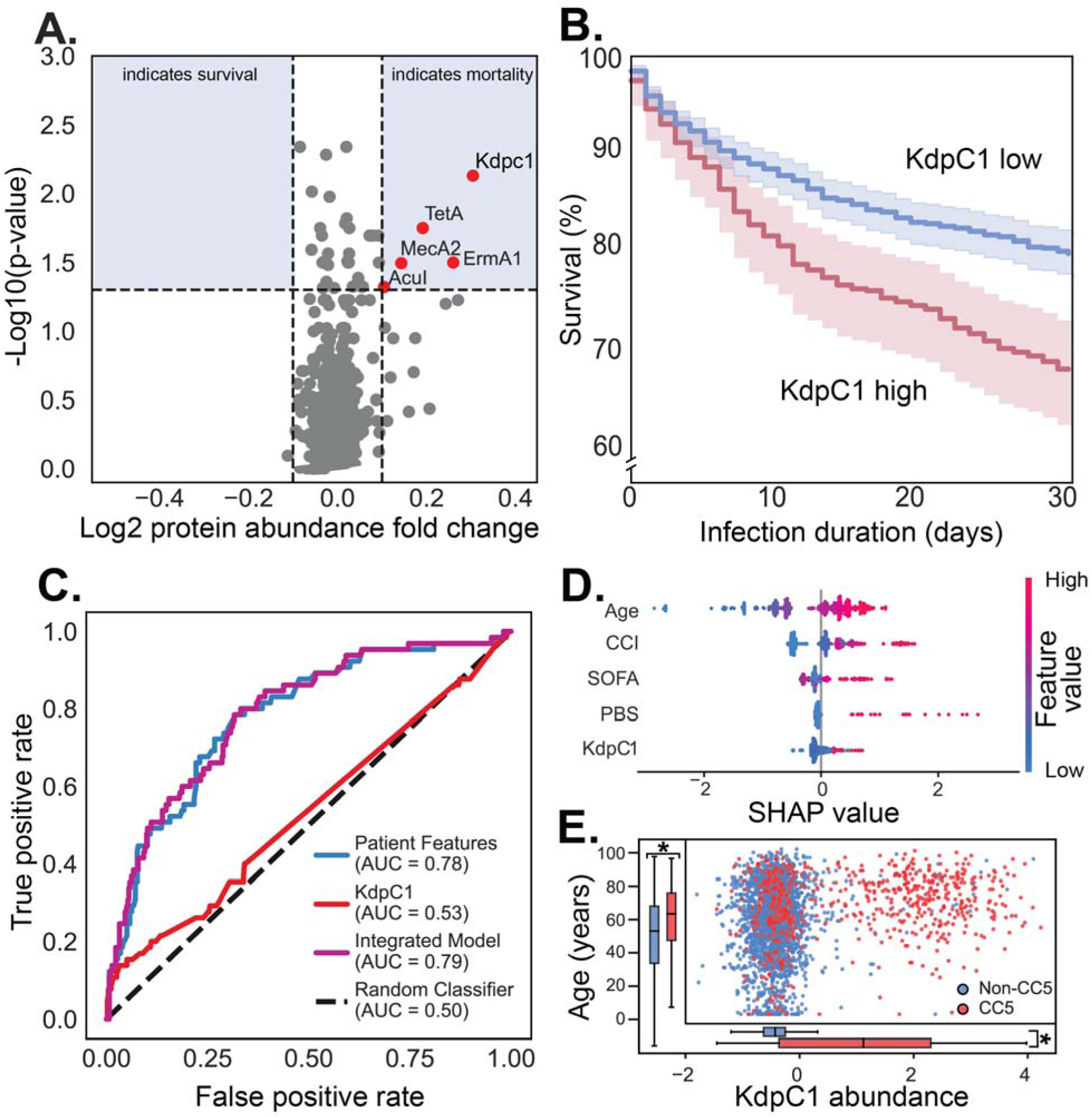
Influence of *S. aureus* microbial protein abundance on 30-day BSI mortality. **(A)** Statistical associations (via Mann-Whitney *U* test with Benjamini-Hochberg false discovery rate correction) between protein abundance and 30-day mortality for BSIs caused by *S*. *aureus*. Red points in the blue shaded region indicate proteins which (1) are significantly associated with mortality (corrected *p*<0.05, horizontal dashed lines), and (2) have an absolute log2 fold change exceeding 0.10 (vertical dashed lines). Log2 protein abundance fold change indicates the difference in mean protein abundance between BSIs that did (positive values) or did not (negative values) result in death within 30 days. **(B)** Percent survival for *S. aureus* BSIs with high or low KdpC1 protein abundance. **(C)** Receiver operating characteristic curves for models designed to predict 30-day BSI patient mortality using a set of four patient features (age, Charlson comorbidity index [CCI], Pitt bacteremia score [PBS], and sequential organ failure assessment [SOFA]), KdpC1 protein abundance, or both. **(D)** SHAP analysis on the integrated patient-and-microbe BSI mortality model. An interpretation guide for this plot is provided in the caption of Figure 2. **(E)** Relationship between patient age, KdpC1 abundance, and *S. aureus* clonal complex (CC). The significance indicator (*) on the inset boxplots denotes *p*<0.001.

Our univariate analyses confirm that certain microbial genes and protein abundance levels co-segregate with patient outcomes. Therefore, we anticipated that including microbial genomic and proteomic data in our ML models would significantly improve our ability to predict outcome. To test this, we constructed ML models of 30-day mortality separately for four common BSI pathogens (*E. coli*, *S. aureus*, *K. pneumoniae*, and *E. faecium*) using the presence/absence of 8,863–68,192 genes (depending on the species) and 1,805–2,672 protein abundance levels (Table S4). Surprisingly, these comprehensive molecular details provided minimal predictive value beyond our patient-only models. When considered by themselves, models built on the molecular traits alone had AUCs ranging from 0.48-0.64 (Table S3 and Fig. S7), which is only slightly better than a random classifier. Likewise, adding these microbial traits to models with EMR features did not substantially improve our predictions of 30-day mortality (relative to patient-only models; Table S3 and Fig. S7). Moreover, this pattern was replicated in models built from sub-cohorts that were matched for age, CCI, and PBS (Table S3).

To verify this unexpected result, we constructed another set of ML models to probe the predictive value of established virulence and antimicrobial resistance traits (see *Analysis of virulence* in methods). We anticipated that these established outcome-linked traits would improve the predictive power of our models. However, we once again found that these microbial traits did not improve mortality predictions beyond our simplified four-factor patient model of 30-day mortality (Table S3). In summary, complex biochemical details of each microbial pathogen, including the presence of established virulence factors, appear to have little importance for predicting mortality relative to patient factors.

To better understand why the microbial risk factors identified through univariate statistical analyses are not strong predictors of outcome in mixed ML models that include patient factors, we analyzed the co-segregation of microbial traits with patient demographics. Two specific examples of traits analyzed include the presence or absence of *papG* in *E. coli* and abundance of KdpC1 in *S. aureus.* These factors were the most significant outcome-linked genes and protein abundance levels in our dataset, as judged by univariate statistical analysis (Figs. 4a-b and 5a-b). Yet again, these outcome-linked traits failed to improve the performance of ML models when patient factors were considered (Figs. 4c-d and 5c-d). In both cases, analyses of patient demographics versus the microbial traits showed that the risk factors were not uniformly distributed across patients. In the case of *papG,* isolates containing the gene were associated with significantly younger patients (*p*<0.001 via Mann-Whitney U test; *n*=10,501) who were more often female (*p*<0.001 via Chi square test; Fig. 4e). In the case of KdpC1, higher protein abundance levels were significantly associated with older patients (*p*<0.001 via Mann-Whitney U test; *n* = 8,078) and almost exclusively occurred in isolates representing clonal complex 5 (CC5) of *S. aureus* (Fig. 5e). In summary, the microbial traits we identified as being significantly linked to outcome via univariate statistical analyses co-segregated with either high-risk or low-risk patient cohorts; therefore, adding microbial traits to our ML models provided minimal additional predictive power.

### Interpretations

The clinical presentation of BSIs is thought to be shaped by a complex interplay of host and microbial factors. However, there is little data that quantifies the magnitude of these contributions to human infections. The primary objective of this study was to provide a concrete benchmark for assessing the relative contributions of host versus microbial factors in predicting mortality. Our strategy was to develop ML models of outcome based on patient factors, then quantify the performance gains achieved by adding complex multi-omics profiles of microbial isolates from each infection to these models.

Although several groups have developed ML models of BSI mortality using EMR data (*36–42*), these models have never been integrated with comprehensive molecular information about the pathogen. Our central hypothesis was that biochemical traits of the microbes, such as virulence factors or resistance traits, may provide the key to unlocking accurate predictions of mortality. However, after evaluating over a million ML models and >70,000 microbial traits, we have come to the surprising conclusion that the molecular details of pathogens contain little unique information for predicting who lives and who dies.

One possible interpretation of our findings is that ML models have flaws that prevent them from accurately predicting clinical outcome from BSIs. However, this interpretation contradicts the literature (*36–42*) and our own EMR-based models (Fig. 2), both of which achieve similar performance benchmarks and identify similar sets of relevant risk factors. Another interpretation is that our dataset does not contain risk-related microbial traits. However, our univariate statistical analyses identified dozens of microbial traits that were significantly linked to 30-day mortality (Figs. 4a-b, 5a-b, S5 and S6), including diverse outcome-linked virulence factors (e.g., *papG*) and resistance traits (e.g., *vanX* and *mecA*) that have been previously reported in the Calgary population (*66–68*). Consequently, we conclude that our ML models are effective for predicting clinical outcome and that the dataset includes the established microbial risk factors.

A more plausible interpretation of our unexpected findings is that microbial traits are not independent of the patient factors in BSIs. Our analyses consistently show age to be one of the most significant contributors to the predictive performance of our models. Given that multiple recent studies have demonstrated age-linked changes in the skin and gut microbiome (*69*, *70*)— and the fact that many BSIs originate from the microbiota (*71*, *72*)—it is intuitive that we observe non-independence of microbial factors and patient age. Moreover, there is significant literature showing that elderly patients are at elevated risk of contracting infections with certain species (e.g., *E. faecium*) and sequence types (e.g., *E. coli* ST131) (*73–77*). This phenomenon contributes to age-related co-variance, which we observed in our ML analysis of microbial species classification (Fig. 3c). Our analysis of microbial biochemical traits appears to be following this same trend, as exemplified by *papG* in *E. coli* and KdpC1 in *S. aureus*. Our univariate statistical analyses showed that both of these traits were linked to significant differences in mortality rates (Figs. 4a-b and 5a-b). However, neither trait improved the predictive performance of ML models that included patient factors. The reason for this is that *papG* and KdpC1 significantly co-segregate with cohorts of high-risk (KdpC1) or low-risk (*papG*) patients. Therefore, *papG* and KdpC1 appear to be serving as proxy reporters for patient risk strata. This co-segregation of patient risk strata and microbial biochemical traits suggests that caution is warranted when interpreting reports of risk-linked microbial traits.

This study has several important limitations that must be considered when formulating broader conclusions about host-versus-microbial risk factors. Firstly, all of the data were derived from the Calgary metropolitan region and thus carry a regional bias in human and microbial diversity. This geographic focus may affect the generalizability of our findings. However, Calgary’s human and microbial diversity is analogous to other cities in North America and Europe, and we therefore anticipate that our core findings will be transferable within these regions. Secondly, all the patients involved in this study had BSIs, and thus there is no outgroup of healthy control patients or non-infectious isolates. As a result, this study cannot identify human or microbial traits that are risk factors for contracting bloodstream infections. Thirdly, all the patients involved in this study were treated in well-resourced hospital settings. Thus, our findings may not translate to resource-limited regions, especially health regions where patients do not have access to effective front-line antimicrobial therapies. Fourthly, we analyzed the molecular profiles of four species of BSI pathogens in this study (representing 61% of all infections in the cohort)—our data do not report on microbially-linked risk factors in the remaining species. Fifthly, protein abundance levels were measured in *in vitro* cultures and therefore may diverge from those observed during infection. Finally, we cannot exclude the possibility that there are cryptic associations between microbes and BSI outcome that are beyond the reach of the ML tools used in this study. Taking these limitations into consideration, we offer the following summary: ***the molecular characteristics of microbes have a minimal impact on a patient’s likelihood of dying after a patient presents with a BSI caused by one of the four species we evaluated, assuming the patient is treated in a well-resourced hospital***.

With regards to future directions, the dominance of patient-related factors in shaping the trajectory of BSIs may provide a path to new clinical tools. We showed that the outcome of most bloodstream infections can be accurately predicted with just four patient related traits (age, CCI, SOFA, and PBS) that are available in critical care settings. To facilitate the assessment and refinement of our simplified model, we created a web portal (www.resistanceDB.org/software) that allows researchers to predict BSI outcomes from these clinical traits. Future improvements to this model may include immune factors, which have a well-established link to BSI mortality (*5*, *43*, *78*).

## Data Availability

All microbial data produced are available online at www.resistanceDB.org

## Acknowledgments

This work was conducted at the Alberta Centre for Advanced Diagnostics at the University of Calgary and computing resources were housed and maintained in part at the Calgary Metabolomics Research Facility, which is supported by the International Microbiome Centre. Microbial genomics data were generated at the Broad Institute of MIT and Harvard in Cambridge, Massachusetts, USA. In addition to the main authors listed in the author byline and the *Author Contributions* section, the work described in this manuscript is a result of the foundational scientific contributions of the PRIME Consortium (www.resistanceDB.org/prime). Additional members of this consortium are listed in the *Consortium Authorship* section below.

## Funding

Large Scale Applied Research Project competition award (2017) from Genome Canada, administered via Genome Alberta (all authors)

Alberta Innovates Translational Health Chair (IAL)

UCalgary Research Excellence Chair (IAL)

Canada Foundation for Innovation, CFI-JELF 34986 (IAL)

CIHR Canada Graduate Scholarship (RG)

Alberta Graduate Excellence Scholarship (RG)

National Institute of Allergy and Infectious Diseases, National Institutes of Health, Department of Health and Human Services, U19AI110818 (AME) and F32AI179151 (JTS)

## Author contributions

Conceptualization: IAL, AME, YHG, DBG, HB, RG,

Methodology: RG, MEVT, AW, AM, MEM, BJW, GR, RS, JTS, TMP, AP, SW, LFP, RAG, TR, AUL, MH, TDM, JMC, BD

Investigation: RG, MEVT, AW, AM, BJW, AP, HB

Validation: RG, MEVT

Software: RG, MEVT, SW

Project Administration: IAL

Funding acquisition: IAL

Supervision: IAL, MEM, AME, YHG

Writing – original draft: RG, IAL

Writing – review & editing: RG, IAL, AME, YHG, HB, DBG, TDM, MEVT, JMC

## Competing interests

The authors declare that they have no competing interests.

## PRIME Consortium

PRIME Consortium members who directly contributed to this study include:

Primary authors

Rory Gilliland^1^, Mario E. Valdés-Tresanco^2^, Andriy Plakhotnyk^2^, Anika Westlund^2^, Joshua T. Smith^3^, Thi Mui Pham^4^, Tatum D. Mortimer^5^, Bruce J. Walker^3^, Ashlee M. Earl^3^, Yonatan H. Grad^4^, M. Ethan MacDonald^1,6–8†^, and Ian A. Lewis^2^*^†^,

Contributing Authors (listed alphabetically)

Hallgrimur Benediktsson^9,10^, John M. Conly^9,11,12^, Bruce Dalton^13^, Daniel B. Gregson^9,11^, Ryan A. Groves^2^, Morgan Hepburn^2^, Alikhan Mansuri^2^, Luis F. Ponce^2^, Gopal Ramamourthy^2^, Thomas Rydzak^2^, Rauf Salamzade^3^, Annegret Ulke-Lemee^2^, and Soren Wacker^2^

^1^Department of Biomedical Engineering, University of Calgary; Calgary, T2N 1N4, Canada

^2^Alberta Centre for Advanced Diagnostics (ACAD), Department of Biological Sciences, University of Calgary; Calgary, T2N 1N4, Canada

^3^Broad Institute of MIT and Harvard; Cambridge, MA, 02142, United States

^4^Harvard T.H. Chan School of Public Health; Boston, MA, 02115, United States

^5^Department of Population Health, University of Georgia; Athens, GA, 30602, United States

^6^Department of Electrical & Software Engineering, University of Calgary; Calgary, T2N 1N4, Canada

^7^Department of Radiology, University of Calgary; Calgary, T2N 1N4, Canada

^8^Hotchkiss Brain Institute, University of Calgary; Calgary, T2N 1N4, Canada

^9^Department of Pathology and Laboratory Medicine, University of Calgary; Calgary, T2N 1N4, Canada

^10^Alberta Precision Laboratories, Calgary, T2L 2K8, Canada

^11^Department of Medicine, Cumming School of Medicine, University of Calgary, Calgary, T2N 1N4, Canada

^12^Infection Prevention and Control, Alberta Health Services, Calgary, Alberta, Canada

^13^Pharmacy Services, Alberta Health Services, Calgary, Alberta, Canada

Additional Members (PRIME consortium members who contributed to the foundational elements of the program)

Sergei Y. Noskov, Herman Barkema, Gregory Tyrrell, Marcello Tonelli, Tanis Dingle, Michael Parkins, Otto G. Vanderkooi, Edward L. Huttlin, Joe Harrison, Diego Nobrega, Nadia Monych, Amy Poelzer, Yeganeh Khaniani, Aja List, Jason Lam, Lindsey Paul, Nick Fitzgerald, Sean English, Jimmy Harold, Matthew Rogers, Owen Conroy, Fiona Clement, Colin MacKenzie, Racheal Apata, Nithia Swaminathan, Rachel Newmiller, Troy D. Feener, Maryam Mapar, Jenna Poelzer, Amy Poelzer, and Madeline Kline. A complete listing of all PRIME Consortium members can be found at www.resistanceDB.org/prime.

## Data and materials availability

Microbial data are available from www.resistanceDB.org.

## Supplementary Materials

Materials and Methods

Figs. S1 to S7

Tables S1 to S5

## Notes

### Competing Interest Statement

The authors have declared no competing interest.

### Author Declarations

The institutional research ethics board of the University of Calgary gave ethical approval for this work.

