## Supplementary material for "Microbial traits have minimal power for predicting death from bloodstream infection": Materials and Methods Figs. S1 to S7 Tables S1 to S5

**The PDF file includes:**

Materials and Methods

Figs. S1 to S7

Tables S1 to S5

Materials and Methods

Data Collection for *Calgary BSI Cohort*

*Patient Data*

The *Calgary BSI Cohort* (*CBSIC)* contains 35,746 microbial isolates from patients hospitalized with bloodstream infections (BSIs) in the Calgary health region between January 2006 and October 2022. Patient electronic medical records (EMR) data were provided by Alberta Health Services databases, including the Discharge Abstract Database <https://cumming.ucalgary.ca/sites/default/files/teams/30/DAD_elements.xlsx>, Vital Statistics <https://cumming.ucalgary.ca/sites/default/files/teams/30/VitalStatisticsDeath_elements.xlsx>, and National Ambulatory Care Reporting System <https://cumming.ucalgary.ca/sites/default/files/teams/30/NACRS_elements.xlsx>. This information consists of patient demographics (*e.g.*, age and sex), clinical laboratory measurements (*e.g.*, blood urea nitrogen, bilirubin, and platelets), administrative records (*e.g.*, date of hospital admission and death, where applicable), and comprehensive reporting of tenth-revision International Classification of Diseases (ICD-10-CA) codes (Table S1). Since laboratory data are collected in multiple batches over time, only the earliest set of lab data linked to each infection was used to generate predictions in our model. Likewise, only ICD-10-CA codes assigned at the point of hospital admission were included. EMR data from the *CBSIC* were used to derive indices of comorbidity and disease severity commonly used in the clinic, including the Charlson Comorbidity Index (CCI) (*1*), Pitt Bacteremia Score (PBS) (*2*, *3*), and Sequential Organ Failure Assessment (SOFA) score (*4*). The collection of data for this study was approved by the University of Calgary Institutional Research Ethics Board under protocol REB18-0233. The use of de-identified, routinely collected clinical and microbiology data involved no direct patient contact nor intervention therefore the review board waived the need written informed consent, in accordance with federal regulations.

*Clinical Microbiology Records and Omics Data*

The *CBSIC*’s EMR data are linked to microbiological records (species classification) provided by Alberta Precision Laboratories (APL) for each isolate in the cohort. Each isolate linked to these records was stored in glycerol at –80 °C. Each isolate was thawed and recultured in Mueller Hinton II broth (VWR, 90003-966) in a 96-well deep-well plate at 37 °C with 5% CO_2_. Cultures were grown to logarithmic phase (average optical density A_600_ ~ 0.4) and complete cultures (microbes and supernatant) were refrozen at –80 °C until further processing.

For genomics analyses, cultures of *S. aureus*, *E. coli*, *E. faecium*, and *K. pneumoniae* were uniformly thawed, and microbes were collected via centrifugation. Cells were lysed, and microbial DNA was purified using the MagMax DNA purification kit (AM1840, Thermo Fisher). DNA was quantified, and whole-genome sequencing was performed at the Broad Institute of MIT and Harvard (Cambridge, MA, US). Sequencing libraries were generated using the Nextera XT DNA library prep kit (Illumina, San Diego, CA, US), and 151x151 bp paired-end sequencing was performed on the Illumina HiSeqX platform. Assembly of genomes, gene annotation, and identification of antimicrobial resistance-associated genes from sequencing reads were performed on the computing infrastructure at the University of Calgary using the seQuoia open-source computational pipeline (<https://github.com/broadinstitute/seQuoia>). Clonal Complex (CC) and Multi-Locus Sequence Typing (MLST) data were derived using species-specific scheme databases from PubMLST (*5*) and BIGSdb-Pasteur (https://bigsdb.pasteur.fr/). Data was also processed into a tabular gene presence (1) and absence (0) format using Roary v 3.13.0 (*6*); paralogs were not split into separate groups.

Proteomics analyses were conducted in-house using the QMAP workflow. Briefly, lysis buffer was added to the complete, thawed cultures of *S. aureus*, *E. coli*, *E. faecium*, and *K. pneumoniae*, and microbes were disrupted via ceramic bead homogenization (OMNI BeadRuptor 96; Kennesaw, GA, US). Intracellular and secreted proteins were processed into peptides using single-pot, solid-phase-enhanced sample preparation (*7*), using a Hamilton Nimbus liquid handler. The peptides were normalized and TMT-labelled. Quantitative proteomics data were subsequently collected via liquid chromatography-mass spectrometry (LC-MS), using an Orbitrap Fusion^TM^ Lumos^TM^ Tribrid^TM^ mass spectrometer equipped with EASY-nLC1200, EasySpray NG ion source and FAIMS (Thermo Fisher Scientific). Data were processed into continuous tabular protein expression values.

Data Curation & Preprocessing

All data curation, processing, and analysis was conducted using Python 3.9 (Centrum voor Wiskunde en Informatica, Amsterdam), with the NumPy (*8*), pandas (*9*), and scikit-learn (*10*) libraries forming the backbone of these methods.

*Target Variable Definition*

To facilitate machine learning (ML) classification analyses, the patient outcome variables were defined as binary targets indicating the occurrence (1) or absence (0) of an outcome. The patient mortality variable was defined as patient death within 30 days of the first blood isolate collection. A long hospital stay was defined as a hospital-stay length (in any unit) of more than two weeks. A costly hospital stay was defined as one in which the Canadian Institute for Health Information resource intensity weight (RIW) of the total hospital stay exceeded 2, indicative of a hospital stay cost that was more than double the average. Many patients in the *CBSIC* were admitted to hospital for other reasons but developed BSIs over the course of their stay. Therefore, RIW and hospital stay length reflect the total burden of each full hospitalization event.

*ICD-10-CA Code Preprocessing*

ICD-10-CA code data was processed into a set of 3,949 binary features, each indicating a code that was (1) or was not (0) assigned at the point of hospital admission for each patient. To reduce excessive granularity among these variables, the codes and their descriptions were used to combine groups of these features into single features (using OR logic) representing broader categories of diseases or other conditions. For example, the 193 codes representing diabetes with various severities and complications were combined into a single feature called “Diabetes”. Additionally, to maintain focus on patient-related details in the subsequent ML analysis, codes indicating an infection (*e.g.*, A41.0: Sepsis due to *Staphylococcus aureus*) or treatments given to the patient (*e.g.*, Z51.5: Encounter for palliative care) were excluded as features for the ML analysis. Finally, ICD-10-CA codes (and combined ICD-10-CA code features) assigned to less than 0.1% of infections in the *CBSIC* were excluded. After processing, 229 ICD-10-CA code features remained.

*Variable Exclusion Criteria*

The clinical index features (and patient age) were cross-correlated. Several scores were strongly correlated (*i.e.*, Pearson correlation > 0.25) because they quantify the same underlying conditions (CCI *vs.* Elixhauser Comorbidity Index, PBS *vs.* qPBS, SOFA *vs.* LODS). In these cases, we retained the more commonly recognized score (*i.e.*, CCI, PBS, SOFA) for interpretability and to reduce multicollinearity. We also screened the ICD-10-CA-related features for correlation. Glasgow Coma Scale (*11*) and arterial blood oxygen partial pressure were excluded as features from the *CBSIC* due to missing data.

*Sample Exclusion Criteria*

*CBSIC* samples representing newborn (0 years of age) or older adult (75+ years of age) patients were excluded to mitigate the impact of notable treatment differences for these groups on the analysis. Additionally, where a patient had multiple BSI records within a 30-day window, all but the first were excluded to eliminate duplicate records of the same infection.

*Missing Values*

Samples lacking values for any patient demographic, ICD-10-CA code, or target variable were excluded. Additionally, any samples lacking genomic data or whose genomic data failed quality control thresholds (Table S4) were removed from the dataset.

Missing values in proteomics data were imputed using the median values of each protein where the corresponding gene was present or absent. Proteomic features that, after partial imputation, were missing in over 90% of samples were also removed to avoid imputation bias. Any remaining missing values were handled as part of the downstream ML analysis.

*Processed Datasets*

Cleaned and preprocessed *CBSIC* datasets for each analysis conducted in this study are summarized in Table S4. Table S1summarizes all patient-related features used in the analysis.

General Machine Learning Approach

The general approach used for the ML analyses conducted in this study is summarized graphically in Figure S2.

A held-out test dataset, representing approximately 20% of the whole dataset, was randomly selected with stratification by the target variable. The remaining 80% was used for training and validation. Patient identification numbers assigned by Alberta Health Services, along with single BSI episodes, were used to ensure that no single patient was represented by different infection episodes in both the testing and training-validation datasets.

The ML pipeline was optimized using the training-validation dataset and a comprehensive grid search over preprocessing steps, feature selection techniques, classification algorithms, and their hyperparameters. Details on the pipeline elements considered in this grid search are provided in Figure S2. Each candidate ML pipeline from the grid search was trained and evaluated on the training-validation dataset using a stratified five-fold cross-validation scheme. Performance on both the training and validation datasets from each cross-validation iteration was assessed using AUC, accuracy, balanced accuracy, F1 score, sensitivity, specificity, and precision. The grid search was parallelized on the University of Calgary’s Advanced Research Computing cluster (https://it.ucalgary.ca/research-computing-services/our-resources/high-performance-computing-hpc/advanced-research-computing-arc.)

For the logistic regression, decision tree, random forest, and XGBoost (*12*) classifiers, class imbalance was handled by upweighting the minority class based on its representation in the training dataset. For the K-nearest neighbors and multilayer perceptron classifiers, this was instead done using the synthetic minority oversampling technique (SMOTE) (*13*). Missing values were handled using median imputation based on the training dataset.

The final model for each target variable was selected based on a comprehensive evaluation of performance during the cross-validation-based grid search. This process involved selecting the model that achieved the highest average AUC in validation (*i.e.*, on the five “validation” instances from cross-validation) while achieving a mean training AUC (*i.e.*, on the five “training” instances from cross-validation) within 0.05 points (a measure to penalize overfitting). In the case of ties or near-ties, preference was given to models that (1) achieved high balanced accuracy in cross-validation, (2) had low standard deviations of performance metrics in cross-validation, and (3) were less complex (*i.e.*, selected fewer features in feature selection, used a simpler classifier, etc.). The final model was then frozen and re-trained on the entire training-validation dataset before evaluating its performance on the held-out test dataset using the same metrics detailed above. This process resulted in one final model for predicting a clinical outcome for each analysis (*e.g.*, all-pathogens mortality using patient features, *E. coli* BSI patient mortality using pathogenic multi-omic features, etc.).

To interrogate the modelled relationships between features and BSI clinical outcomes, a model explainability analysis was conducted using SHapley Additive exPlanations (SHAP) (*14*, *15*) on the final models’ testing predictions.

Patient Trait Analysis

*Machine Learning Analysis*

To predict BSI clinical outcomes using patient traits alone, the above-described ML approach was applied to generate a classification model for predicting the (1) patient mortality (binary), (2) hospital stay duration (thresholded binary), and (3) healthcare cost target variables (thresholded binary)￼. BSIs caused by any pathogen represented in the *CBSIC* were considered. Only patient-related traits such as demographics and comorbidities were used as features in this analysis. The details and performance of these three patient trait models are provided in Table S2.

*Simplified Models*

Based on the results of the full-scale patient trait ML analysis, a compact model for each clinical outcome consisting of (1) patient mortality, (2) hospital stay duration, and (3) healthcare cost target variables, was designed to formalize a clinically feasible patient-factor-only BSI diagnostic algorithm. To build and select these models, the same ML approach described above was applied. However, for this implementation, features were selected manually based on (1) ease of measurement in a clinic (*e.g.*, patient demographics were given preference over blood biochemical measurements) and (2) predictive value evidenced by the results of the full-feature model explainability analysis. The performance of these simplified clinical outcome models was then visualized and compared to their full-feature counterparts. The details and performance of the three simplified models are documented in Table S2.

Microbe Trait Analysis

Predictive models and statistical analyses related to microbial molecular characteristics focussed solely on 30-day mortality rates as an outcome.

*Univariate Statistical Analysis*

The relationship of microbial proteomic and genomic features with 30-day patient mortality was first analyzed using traditional statistical methods. The Mann-Whitney *U* test was used to compare protein abundance levels between BSIs that resulted in patient death and those that did not, while the Chi-square test was used to compare proportions of genes present. A Benjamini-Hochberg false discovery rate multiple hypothesis correction was applied to adjust the *p*-values from both tests. Adjusted *p*-values lower than 0.05 were considered significant.

*Machine Learning Analysis*

First, to evaluate the predictive value of species classification as a feature, an all-pathogens machine learning analysis was performed. Three models were created using the previously described machine learning method to predict 30-day mortality in BSI patients: one based solely on patient features, one based solely on pathogen species classification, and one that combines both feature sets. Species classification data were processed into 16 binary species indicators using one-hot encoding.

Similarly, the predictive value of microbial multi-omics features was evaluated through a pathogen-specific analysis with datasets for *E. coli*, *S. aureus*, *E. faecium*, and *K. pneumoniae*. For each pathogen, three additional models were developed: one based solely on patient features, one based only on microbial multi-omics and strain-related features, and one that incorporates all available features.

The performance of the three model types (patient, microbe, and integrated) for each analysis was then compared to evaluate the predictive power of microbial features for BSI patient mortality, both independently and alongside patient markers. Additionally, explainability analysis was performed as needed to explore the relationships between the modeled features and the targets.

*Cohort Machine Learning Analysis*

To assess whether the predictive value of microbial traits differed across subgroups of BSI patients, the machine-learning analyses were repeated in four smaller cohorts. These cohorts were defined based on: (1) constraining patient age, CCI, and PBS within the smallest possible window; (2) maintaining an adequate sample size and frequency of 30-day patient mortality for the ML analysis; and (3) sampling cohorts representing both old and young patients. These cohorts were:

1.	*E. coli* BSI patients with ages 50–69, CCI of 0 or 1, and PBS of 0 or 1 (*n*=1,165)

2.	*S. aureus* BSI patients with ages 50–69, CCI of 0 or 1, and PBS of 0 or 1 (*n*=744)

3.	All-pathogen BSI patients with ages 55–64, CCI of 0, and PBS of 0 (*n*=1,344)

4.	All-pathogen BSI patients with ages 20–39, CCI of 0, and PBS of 0 (*n*=1,548)

Analysis of cohorts 1 and 2 involved microbial multi-omics data, while cohorts 3 and 4 used species classification data. For each cohort, the described ML modelling protocol was applied to select, train, and evaluate models aimed at predicting 30-day BSI patient mortality based on patient features, microbial features, or both. The performance of these three model types (patient, microbe, and integrated) was then compared to determine whether microbial traits provided additional predictive value for clinical outcomes in more homogeneous patient groups.

*Analysis of virulence*

To better understand the impact that microbial virulence traits have on patient outcome, we conducted ML analyses using the following known resistance/virulence factors: the VanX protein in *E. faecium* (*16*), the KdpC protein in *S. aureus*, the *papG* gene in *E. coli* (*17*), the *K. pneumoniae* virulence score defined by Lam and colleagues (*18*), and all *S. aureus* virulence proteins documented in the Virulence Factor Database (*19*). We then constructed ML models to predict 30-day BSI patient mortality using (1) patient features only, (2) the specific microbial feature(s) only, or (3) a combination of the two. The above-described ML protocol was used for this purpose with two key modifications for simplicity. First, the feature selection step was omitted from the grid search (except for in the *S. aureus* virulence factor analysis). Secondly, all patient factors were excluded except for age, CCI, PBS, and SOFA.

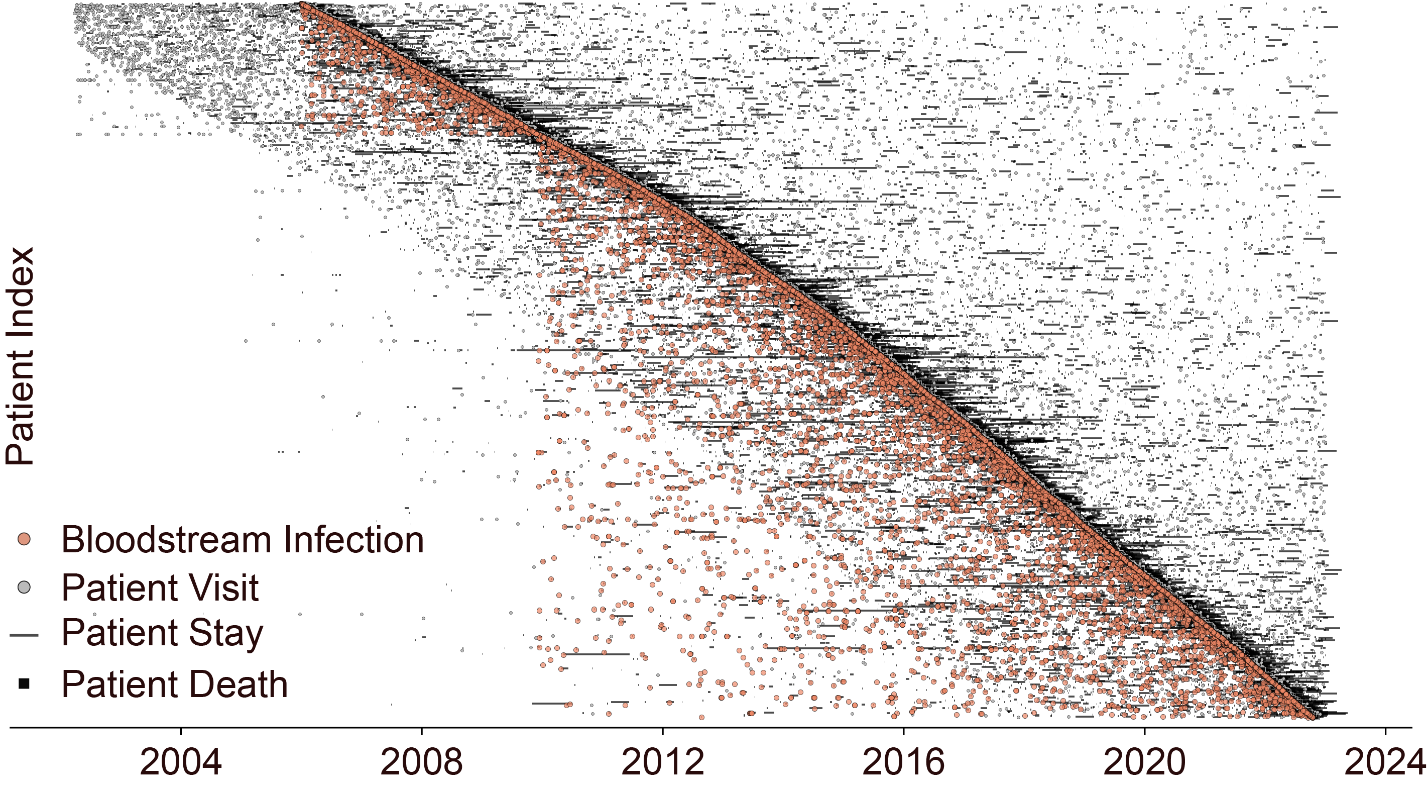

Fig S1. Harmonized clinical microbiology dataset comprising 442,251 patient diagnostic and procedure records linked to 35,746 bloodstream infection isolates and 60,402 patient visits/stays, including 30-day mortality.

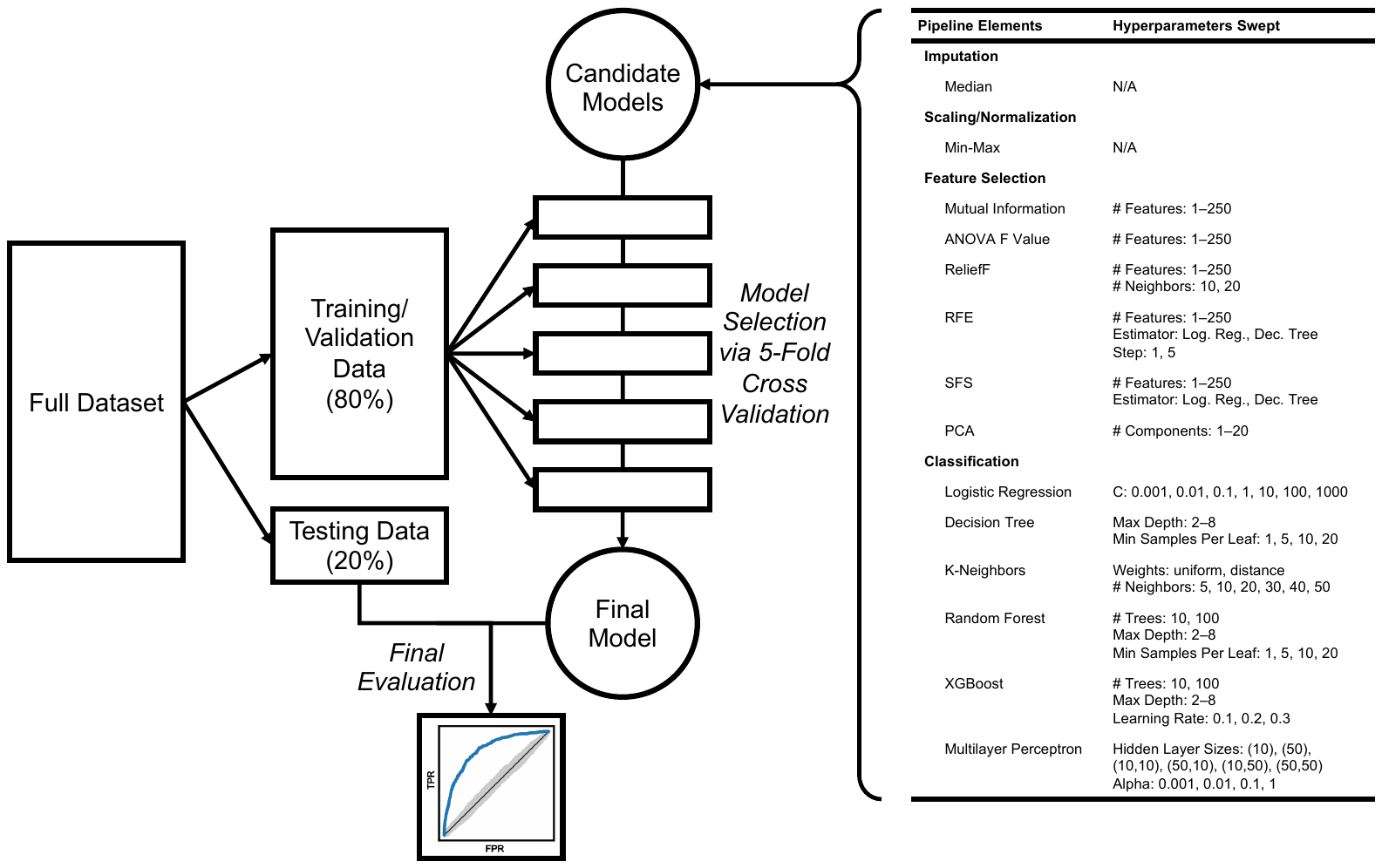

Fig S2. A schematic of the machine learning experimental protocol used in the study to compare candidate models using a cross-validation grid search and evaluate the final model on unseen testing data. The final model is trained on the Training/Validation Data before Final Evaluation. Abbreviations: MRMR, minimum redundancy maximum relevance; RFE, recursive feature elimination; SFS, sequential feature selection; PCA, principal component analysis.

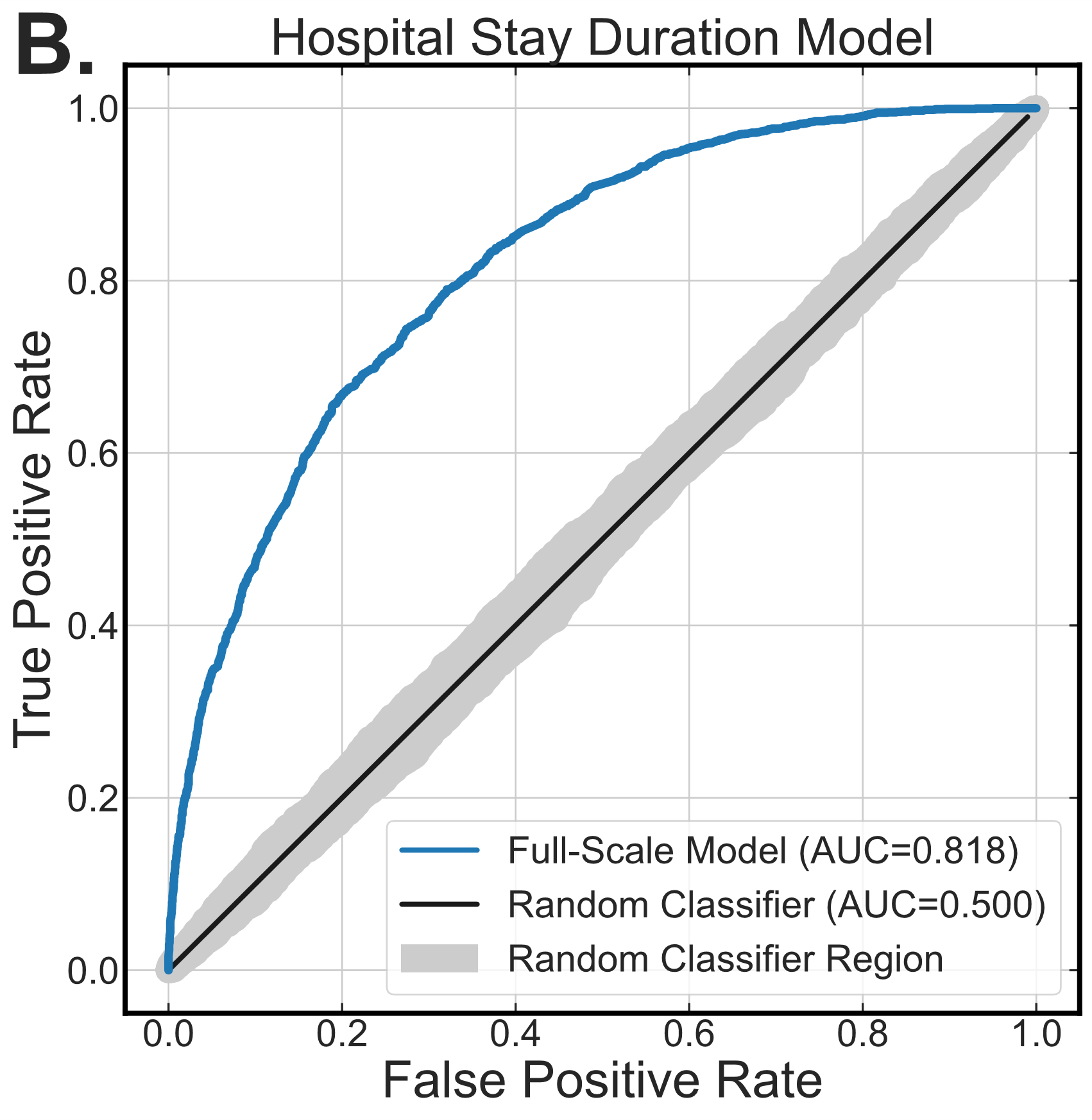

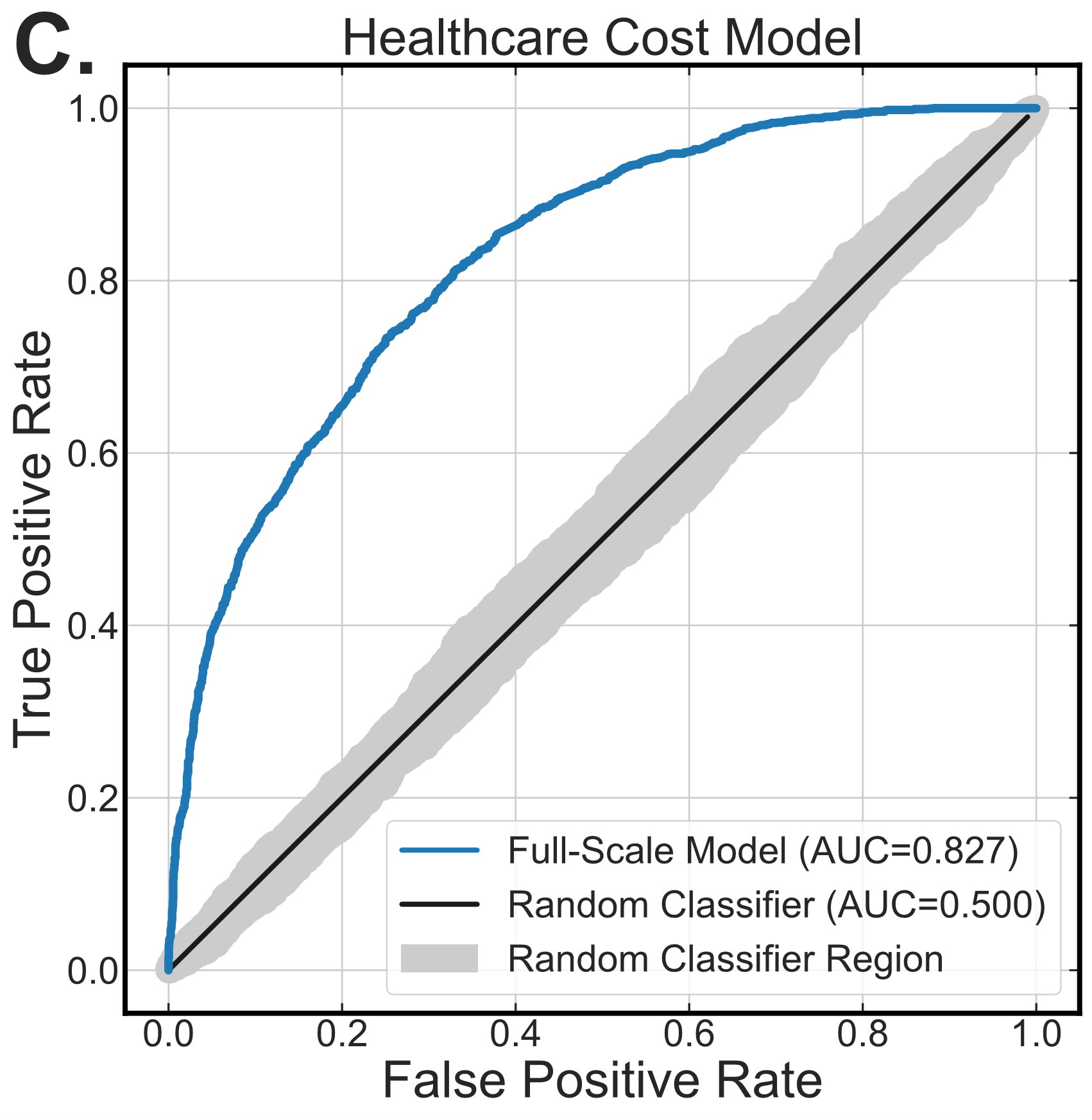

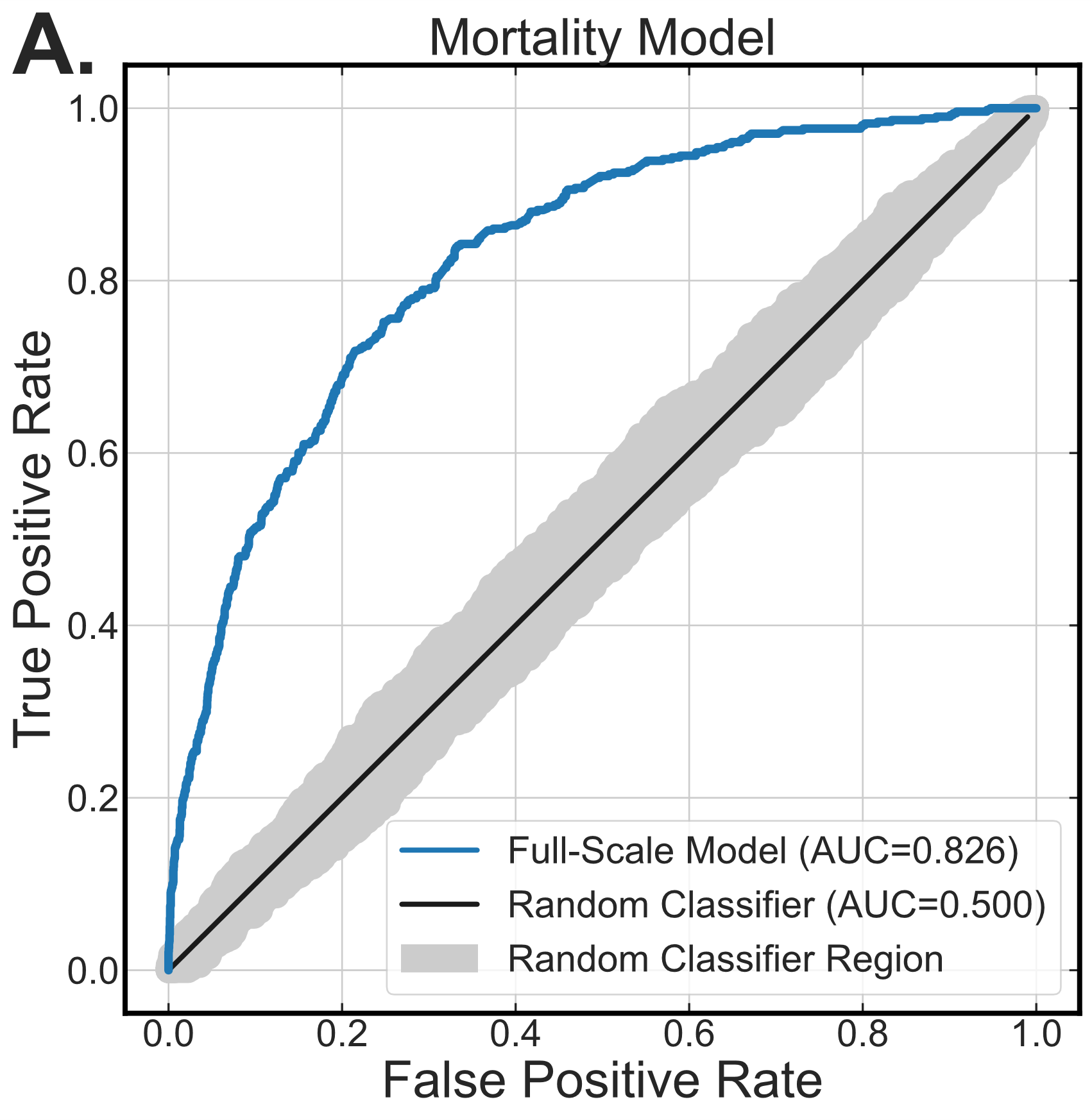

Fig S3. Receiver operating characteristic curves for full-scale patient feature-only clinical outcome prediction models for (A) 30-day mortality, (B) 2-week hospital stay duration, and (C) doubled healthcare cost relative to an average hospital stay for bloodstream infection patients. Abbreviation: AUC, area under the curve.

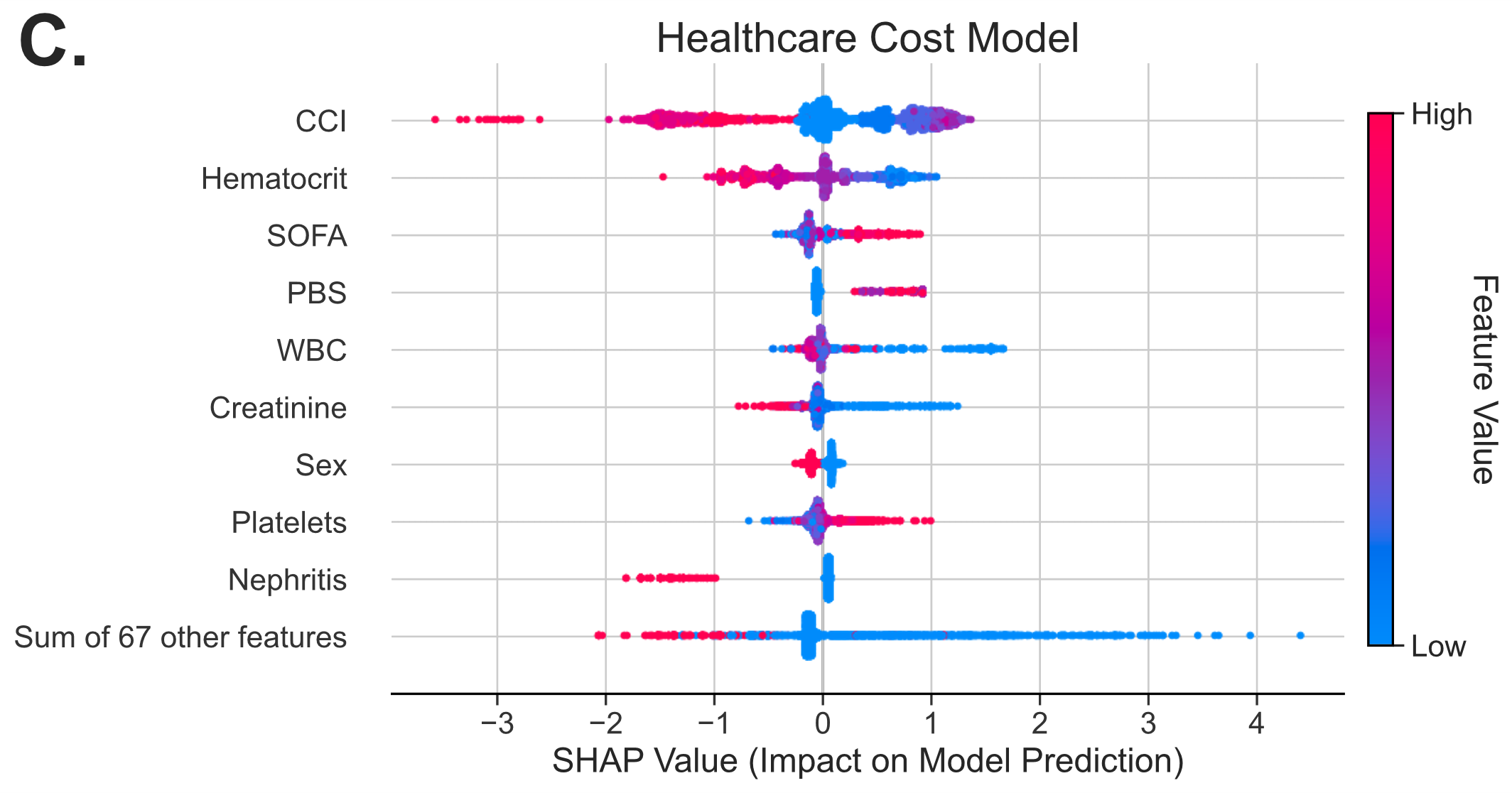

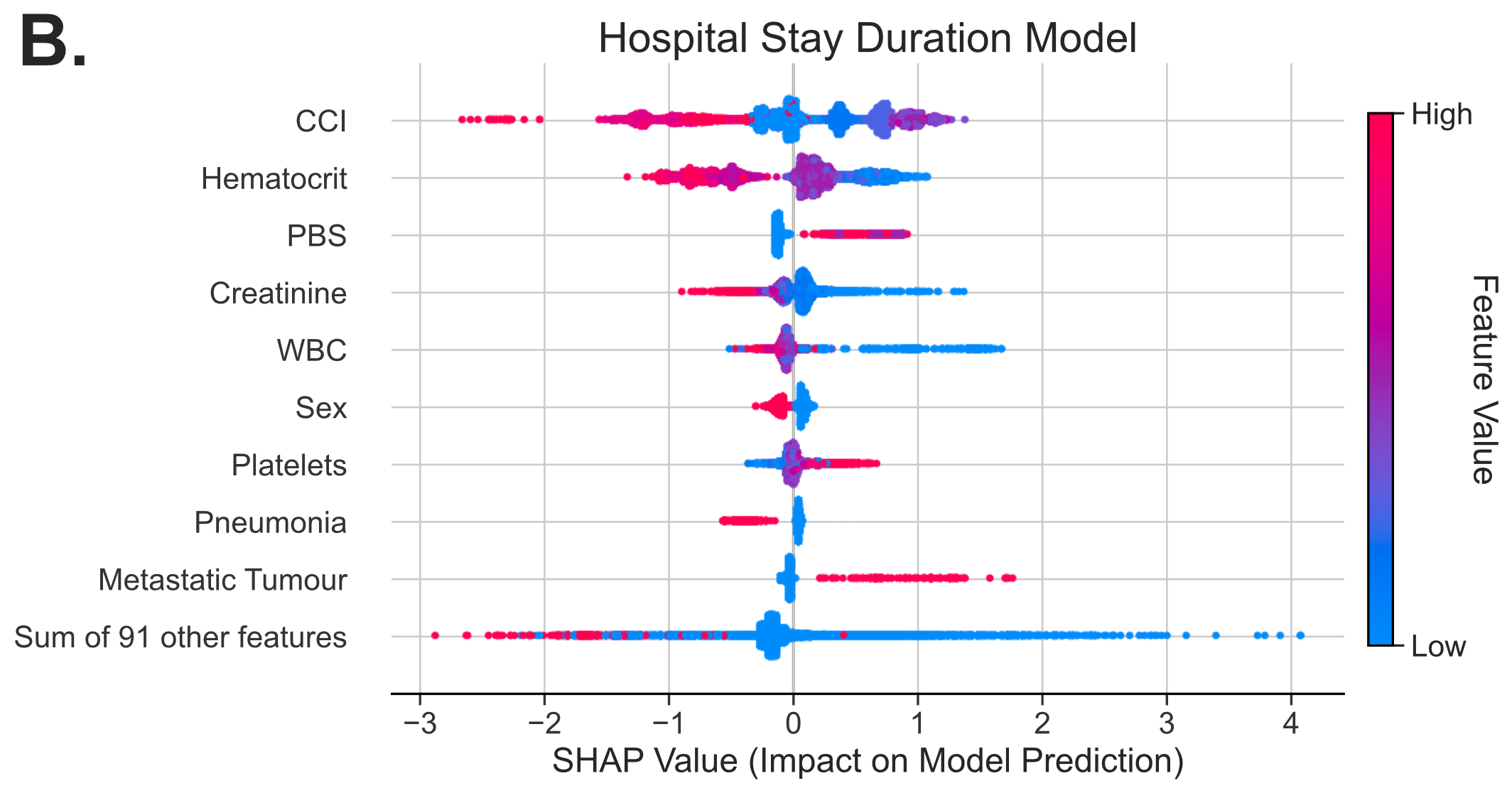

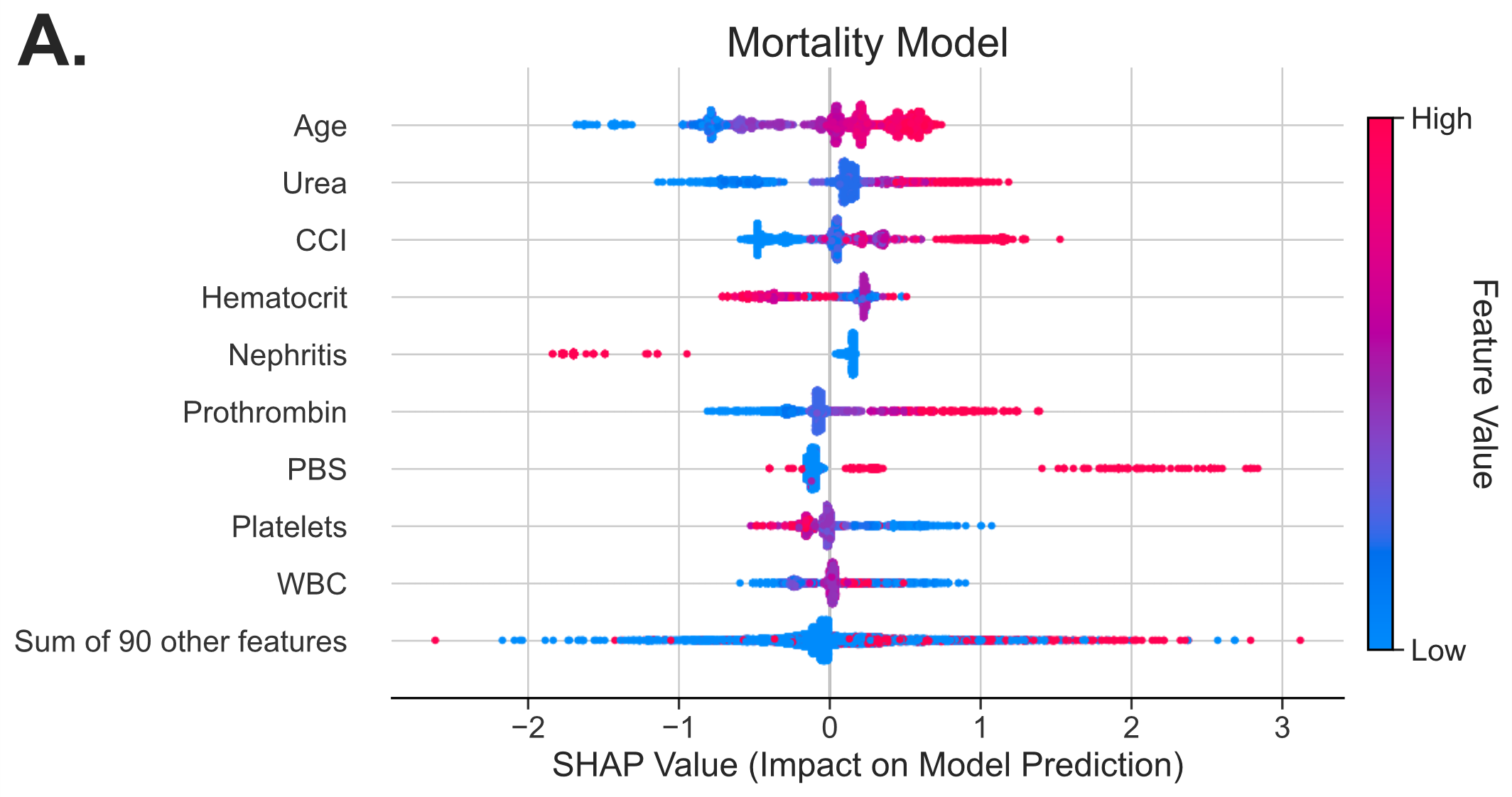

Fig S4. SHAP analysis on the held-out testing dataset predictions of full-scale patient feature-only clinical outcome models for (A) 30-day mortality, (B) 2-week hospital stay duration, and (C) doubled healthcare cost relative to an average hospital stay for bloodstream infection patients. Each dot represents a sample in the held-out testing set and is colored by the value of the corresponding feature in that sample. Dots are assigned SHAP values according to their impact on the model’s prediction, where positive SHAP values denote a sample for which the corresponding feature value was interpreted to increase the risk of an adverse clinical outcome by the model, and vice versa. Note that female patient sex was encoded as 1 (red point) and male as 0 (blue point). Abbreviations: CCI, Charlson comorbidity index; PBS, Pitt bacteremia score; WBC, white blood cell count; SOFA, sequential organ failure assessment. Feature value corresponds to a high value (red) for a given feature (*i.e.*, old age) or a low value (blue) for that same feature (*i.e.*, young age).

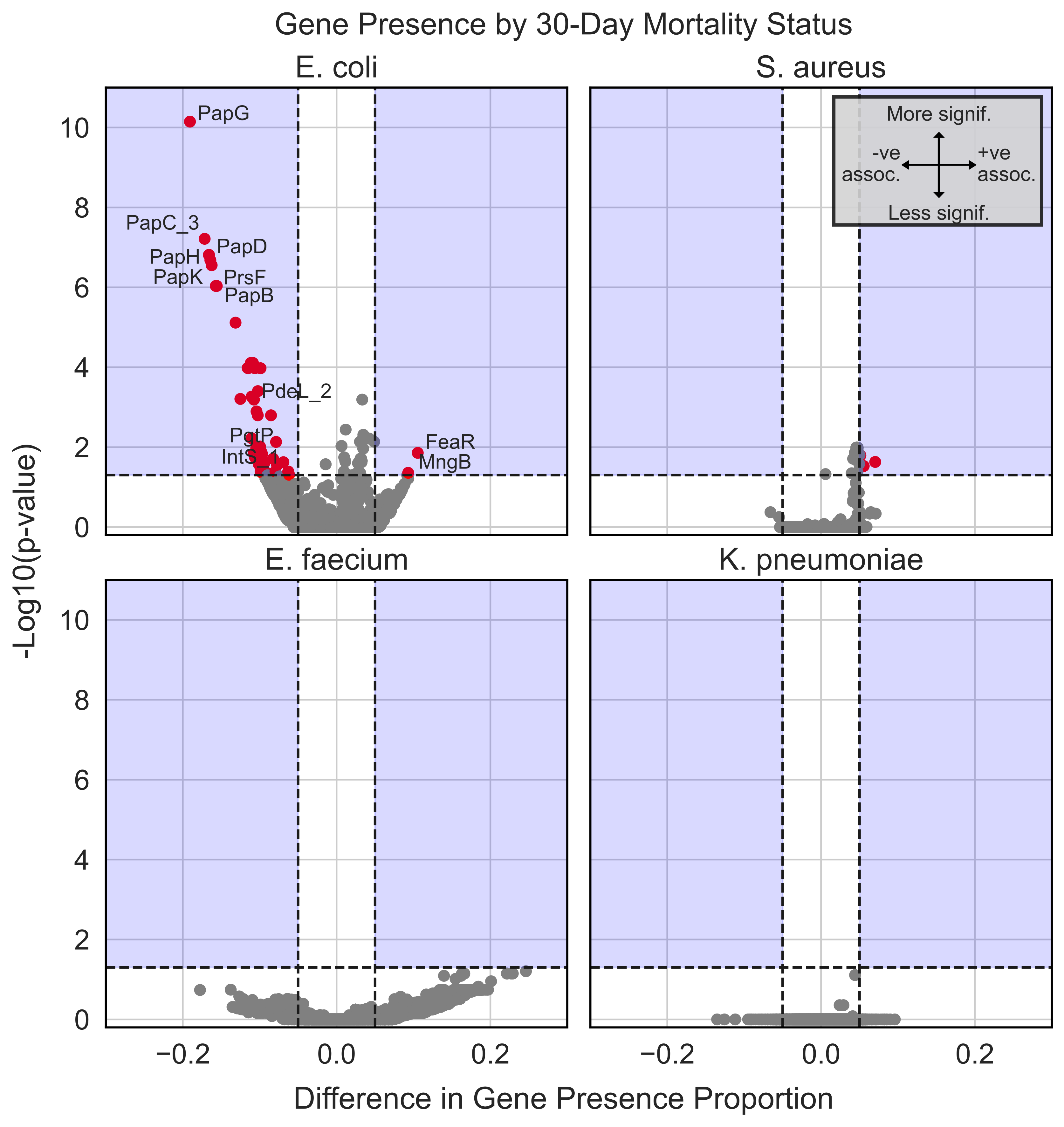

Fig S5. Statistical associations (via Chi-square test with false discovery rate correction) between gene presence and 30-day bloodstream infection (BSI) patient mortality for BSIs caused by *Escherichia coli*, *Staphylococcus aureus*, *Enterococcus faecium*, and *Klebsiella pneumoniae*. Red points in the blue shaded region indicate genes which (1) are significantly associated with mortality (*p*<0.05, horizontal dashed lines) and (2) have an absolute difference in gene presence proportion exceeding 0.05 (vertical dashed lines). A positive difference in gene presence proportion indicates a gene whose presence is enriched in BSIs causing death within 30 days, and vice versa. Some red points are not labelled either for clarity or because the corresponding gene has not yet been named.

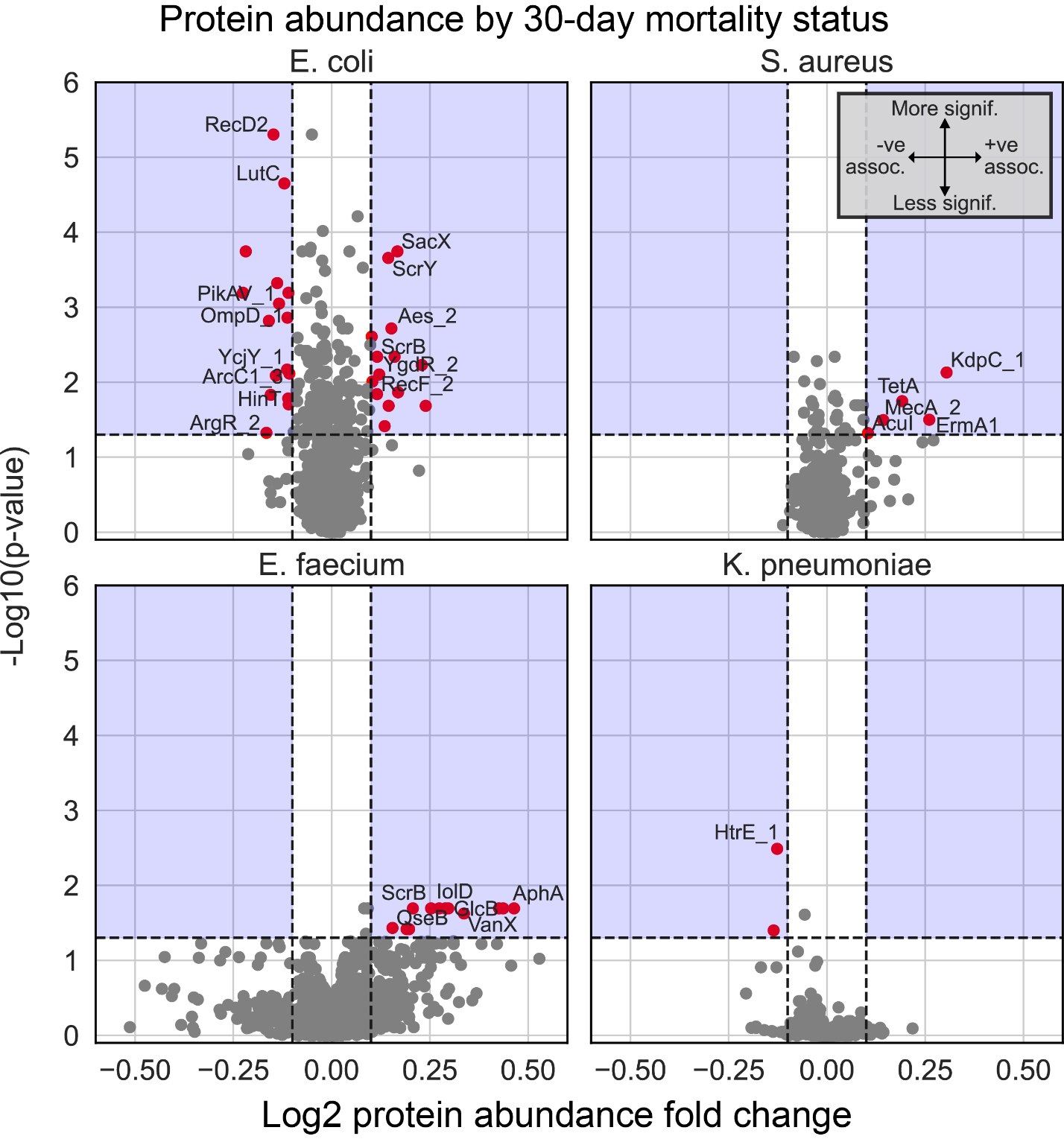

Fig S6. Statistical associations (via Mann-Whitney *U* test with Benjamini-Hochberg correction for multiple hypothesis testing) between protein abundance and 30-day BSI patient mortality for BSIs caused by *E*. *coli*, *S*. *aureus*, *E*. *faecium*, and *K*. *pneumoniae*. Red points in the blue shaded region indicate proteins which (1) are significantly associated with mortality (*p*<0.05, horizontal dashed lines) and (2) have an absolute log2 fold change exceeding 0.10 (vertical dashed lines). Log2 protein abundance fold change indicates the difference in mean protein abundance between BSIs that did and did not result in 30-day mortality, where positive values denote enrichment of a protein in BSIs resulting in mortality. Some red points are not labelled either for clarity or because the corresponding protein was not given a unique name during annotation and pan genome analysis.

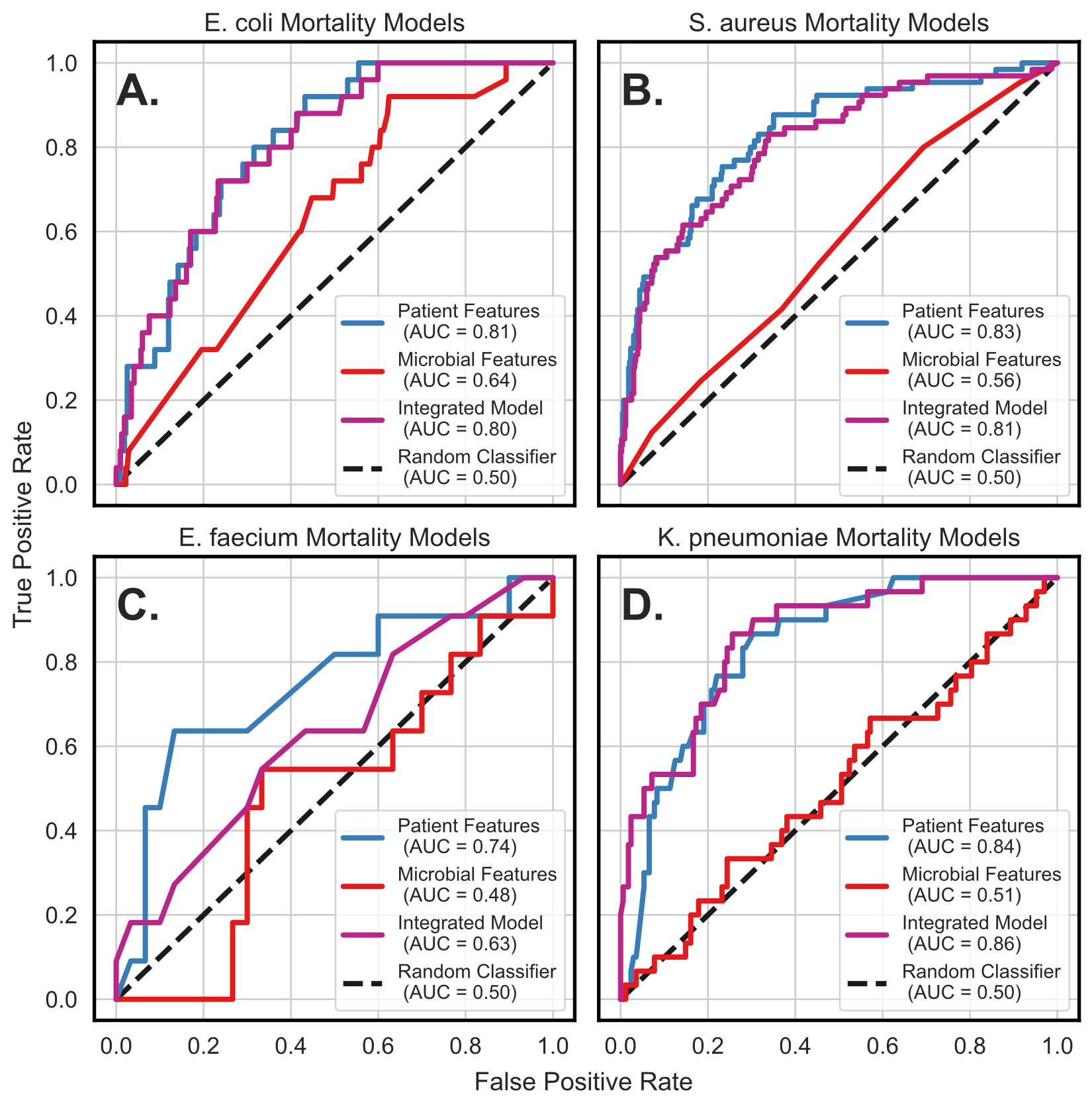

Fig S7. Performances (on held-out test dataset) of patient mortality models developed for (A) *E. coli*, (B) *S. aureus*, (C) *E. faecium*, and (D) *K. pneumoniae* bloodstream infections. Models used patient factors, microbial factors, or both.

Table S1. List of all patient-related features and outcome metrics used in the analyses.

| **Features** | **Description** |
| --- | --- |
| BI_NBR | Unique identifier of blood isolate |
| APL: PID | Unique identifier of patient |
| APL: INDEX_30DAYS | Indicates index isolates |
| AHS: GENDER | Patient sex |
| AHS: NAGE_YR | Patient age in years |
| OUTCOME: DAYS_TO_DEATH | Number of days after blood isolate when patient was deceased, if applicable |
| OUTCOME: ThirtyDay_Death | Indicates patient death on or before 30 days after blood isolate was collected |
| OUTCOME: DAYS_HOSP_AC | Number of days patient was in hospital associated with current blood isolate for any cause |
| OUTCOME: RIW_AC | RIW for current hospital stay |
| COMORB: Charlson_WIC | Charlson Comorbidity Index |
| S.SCORE: PBS | Pitt Bacteremia Score |
| S.SCORE: CURB65 | CURB-65 Score |
| S.SCORE: SOFA | Systematic Organ Failure Assessment score |
| Fever | Indicates patient with a fever |
| Hypotension | Indicates patient with hypotension |
| Ventilation | Indicates patient on ventilatory support |
| GCS | Glasgow Coma Scale |
| Urea | Blood urea nitrogen |
| Tachypnea | Indicates patient with tachypnea |
| PAO2 | Arterial oxygen partial pressure |
| Platelets | Platelet count |
| Bilirubin | Bilirubin |
| Creatinine | Serum creatinine |
| Tachycardia | Indicates patient with tachycardia |
| Bradycardia | Indicates patient with bradycardia |
| WBC | White blood cell count |
| Prothrombin | Prothrombin time |
| Hematocrit | Hematocrit |
| DX: E860 | Indicates assignment of specified ICD-10 code |
| DX: E871 | Indicates assignment of specified ICD-10 code |
| DX: I500 | Indicates assignment of specified ICD-10 code |
| DX: E876 | Indicates assignment of specified ICD-10 code |
| DX: E872 | Indicates assignment of specified ICD-10 code |
| DX: R4180 | Indicates assignment of specified ICD-10 code |
| DX: R18 | Indicates assignment of specified ICD-10 code |
| DX: E875 | Indicates assignment of specified ICD-10 code |
| DX: R090 | Indicates assignment of specified ICD-10 code |
| DX: I330 | Indicates assignment of specified ICD-10 code |
| DX: K650 | Indicates assignment of specified ICD-10 code |
| DX: E870 | Indicates assignment of specified ICD-10 code |
| DX: R64 | Indicates assignment of specified ICD-10 code |
| DX: E868 | Indicates assignment of specified ICD-10 code |
| DX: K831 | Indicates assignment of specified ICD-10 code |
| DX: I519 | Indicates assignment of specified ICD-10 code |
| DX: K8302 | Indicates assignment of specified ICD-10 code |
| DX: J869 | Indicates assignment of specified ICD-10 code |
| DX: R33 | Indicates assignment of specified ICD-10 code |
| DX: K566 | Indicates assignment of specified ICD-10 code |
| DX: J80 | Indicates assignment of specified ICD-10 code |
| DX: K830 | Indicates assignment of specified ICD-10 code |
| DX: F419 | Indicates assignment of specified ICD-10 code |
| DX: K8308 | Indicates assignment of specified ICD-10 code |
| DX: R113 | Indicates assignment of specified ICD-10 code |
| DX: R53 | Indicates assignment of specified ICD-10 code |
| DX: E46 | Indicates assignment of specified ICD-10 code |
| DX: R060 | Indicates assignment of specified ICD-10 code |
| DX: R73812 | Indicates assignment of specified ICD-10 code |
| DX: R410 | Indicates assignment of specified ICD-10 code |
| DX: K590 | Indicates assignment of specified ICD-10 code |
| DX: R138 | Indicates assignment of specified ICD-10 code |
| DX: N200 | Indicates assignment of specified ICD-10 code |
| DX: N419 | Indicates assignment of specified ICD-10 code |
| DX: F432 | Indicates assignment of specified ICD-10 code |
| DX: G934 | Indicates assignment of specified ICD-10 code |
| DX: G039 | Indicates assignment of specified ICD-10 code |
| DX: E877 | Indicates assignment of specified ICD-10 code |
| DX: M6289 | Indicates assignment of specified ICD-10 code |
| DX: N201 | Indicates assignment of specified ICD-10 code |
| DX: J9810 | Indicates assignment of specified ICD-10 code |
| DX: K631 | Indicates assignment of specified ICD-10 code |
| DX: N136 | Indicates assignment of specified ICD-10 code |
| DX: K921 | Indicates assignment of specified ICD-10 code |
| DX: R748 | Indicates assignment of specified ICD-10 code |
| DX: M6280 | Indicates assignment of specified ICD-10 code |
| DX: K550 | Indicates assignment of specified ICD-10 code |
| DX: K20 | Indicates assignment of specified ICD-10 code |
| DX: K920 | Indicates assignment of specified ICD-10 code |
| DX: M549 | Indicates assignment of specified ICD-10 code |
| DX: K660 | Indicates assignment of specified ICD-10 code |
| DX: R55 | Indicates assignment of specified ICD-10 code |
| DX: R21 | Indicates assignment of specified ICD-10 code |
| DX: R51 | Indicates assignment of specified ICD-10 code |
| DX: R5688 | Indicates assignment of specified ICD-10 code |
| DX: Z911 | Indicates assignment of specified ICD-10 code |
| DX: G931 | Indicates assignment of specified ICD-10 code |
| DX: K567 | Indicates assignment of specified ICD-10 code |
| DX: J984 | Indicates assignment of specified ICD-10 code |
| DX: N40 | Indicates assignment of specified ICD-10 code |
| DX: E162 | Indicates assignment of specified ICD-10 code |
| DX: T86001 | Indicates assignment of specified ICD-10 code |
| DX: K767 | Indicates assignment of specified ICD-10 code |
| DX: B24 | Indicates assignment of specified ICD-10 code |
| DX: N180 | Indicates assignment of specified ICD-10 code |
| DX: I81 | Indicates assignment of specified ICD-10 code |
| DX: K559 | Indicates assignment of specified ICD-10 code |
| DX: E878 | Indicates assignment of specified ICD-10 code |
| DX: F411 | Indicates assignment of specified ICD-10 code |
| DX: E222 | Indicates assignment of specified ICD-10 code |
| DX: K659 | Indicates assignment of specified ICD-10 code |
| DX: R4188 | Indicates assignment of specified ICD-10 code |
| DX: R296 | Indicates assignment of specified ICD-10 code |
| DX: R739 | Indicates assignment of specified ICD-10 code |
| DX: R4029 | Indicates assignment of specified ICD-10 code |
| DX: R112 | Indicates assignment of specified ICD-10 code |
| DX: K297 | Indicates assignment of specified ICD-10 code |
| DX: G629 | Indicates assignment of specified ICD-10 code |
| DX: R042 | Indicates assignment of specified ICD-10 code |
| DX: J988 | Indicates assignment of specified ICD-10 code |
| DX: N23 | Indicates assignment of specified ICD-10 code |
| DX: E873 | Indicates assignment of specified ICD-10 code |
| DX: K3188 | Indicates assignment of specified ICD-10 code |
| DX: I4891 | Indicates assignment of specified ICD-10 code |
| DX: F067 | Indicates assignment of specified ICD-10 code |
| DX: W80 | Indicates assignment of specified ICD-10 code |
| DX: G632 | Indicates assignment of specified ICD-10 code |
| DX: K658 | Indicates assignment of specified ICD-10 code |
| DX: N319 | Indicates assignment of specified ICD-10 code |
| DX: I248 | Indicates assignment of specified ICD-10 code |
| DX: K863 | Indicates assignment of specified ICD-10 code |
| DX: K760 | Indicates assignment of specified ICD-10 code |
| DX: R040 | Indicates assignment of specified ICD-10 code |
| DX: K635 | Indicates assignment of specified ICD-10 code |
| DX: K632 | Indicates assignment of specified ICD-10 code |
| DX: N139 | Indicates assignment of specified ICD-10 code |
| DX: R470 | Indicates assignment of specified ICD-10 code |
| DX: F39 | Indicates assignment of specified ICD-10 code |
| DX: K768 | Indicates assignment of specified ICD-10 code |
| DX: B259 | Indicates assignment of specified ICD-10 code |
| DX: R02 | Indicates assignment of specified ICD-10 code |
| DX: W19 | Indicates assignment of specified ICD-10 code |
| DX: I313 | Indicates assignment of specified ICD-10 code |
| DX: R111 | Indicates assignment of specified ICD-10 code |
| DX: K565 | Indicates assignment of specified ICD-10 code |
| DX: J850 | Indicates assignment of specified ICD-10 code |
| DX: K029 | Indicates assignment of specified ICD-10 code |
| DX: G312 | Indicates assignment of specified ICD-10 code |
| DX: I249 | Indicates assignment of specified ICD-10 code |
| DX: R05 | Indicates assignment of specified ICD-10 code |
| DX: R591 | Indicates assignment of specified ICD-10 code |
| DX: I499 | Indicates assignment of specified ICD-10 code |
| DX: G20 | Indicates assignment of specified ICD-10 code |
| DX: K298 | Indicates assignment of specified ICD-10 code |
| DX: N210 | Indicates assignment of specified ICD-10 code |
| DX: R740 | Indicates assignment of specified ICD-10 code |
| DX: K912 | Indicates assignment of specified ICD-10 code |
| DX: H669 | Indicates assignment of specified ICD-10 code |
| DX: O60101 | Indicates assignment of specified ICD-10 code |
| DX: I200 | Indicates assignment of specified ICD-10 code |
| DX: S22500 | Indicates assignment of specified ICD-10 code |
| DX: R068 | Indicates assignment of specified ICD-10 code |
| DX: I509 | Indicates assignment of specified ICD-10 code |
| DX: K219 | Indicates assignment of specified ICD-10 code |
| DX: F603 | Indicates assignment of specified ICD-10 code |
| DX: I4900 | Indicates assignment of specified ICD-10 code |
| DX: R392 | Indicates assignment of specified ICD-10 code |
| DX: I864 | Indicates assignment of specified ICD-10 code |
| DX: R628 | Indicates assignment of specified ICD-10 code |
| DX: I319 | Indicates assignment of specified ICD-10 code |
| DX: E274 | Indicates assignment of specified ICD-10 code |
| DX: H050 | Indicates assignment of specified ICD-10 code |
| DX: Z722 | Indicates assignment of specified ICD-10 code |
| DX: Z602 | Indicates assignment of specified ICD-10 code |
| DX: U071 | Indicates assignment of specified ICD-10 code |
| DX: F319 | Indicates assignment of specified ICD-10 code |
| DX: N280 | Indicates assignment of specified ICD-10 code |
| DX: M2546 | Indicates assignment of specified ICD-10 code |
| DX: G439 | Indicates assignment of specified ICD-10 code |
| DX: TUBERCULOSIS | Indicates assignment of specified processed ICD-10 code |
| DX: HERPES | Indicates assignment of specified processed ICD-10 code |
| DX: HEPATITIS | Indicates assignment of specified processed ICD-10 code |
| DX: PEDICULOSIS | Indicates assignment of specified processed ICD-10 code |
| DX: HYPOTHYROIDISM | Indicates assignment of specified processed ICD-10 code |
| DX: DIABETES | Indicates assignment of specified processed ICD-10 code |
| DX: METABOLISM DISORDERS | Indicates assignment of specified processed ICD-10 code |
| DX: ENCEPHALITIS, MYELITIS, AND ENCEPHALOMYELITIS | Indicates assignment of specified processed ICD-10 code |
| DX: EPILEPTIC DISORDERS | Indicates assignment of specified processed ICD-10 code |
| DX: SLEEP | Indicates assignment of specified processed ICD-10 code |
| DX: -PLEGIA | Indicates assignment of specified processed ICD-10 code |
| DX: VALVULAR DISORDER | Indicates assignment of specified processed ICD-10 code |
| DX: ATRIOVENTRICULAR BLOCK | Indicates assignment of specified processed ICD-10 code |
| DX: HYPERTENSIVE DISEASE | Indicates assignment of specified processed ICD-10 code |
| DX: MYOCARDIAL INFARCTION | Indicates assignment of specified processed ICD-10 code |
| DX: ATHEROSCLEROTIC DISEASE | Indicates assignment of specified processed ICD-10 code |
| DX: PERICARDITIS | Indicates assignment of specified processed ICD-10 code |
| DX: STROKE | Indicates assignment of specified processed ICD-10 code |
| DX: ANEURYSM | Indicates assignment of specified processed ICD-10 code |
| DX: EMBOLISM AND THROMBOSIS | Indicates assignment of specified processed ICD-10 code |
| DX: VARICOSE VEINS | Indicates assignment of specified processed ICD-10 code |
| DX: HAEMORRHOIDS | Indicates assignment of specified processed ICD-10 code |
| DX: HYPOTENSION | Indicates assignment of specified processed ICD-10 code |
| DX: SINUSITIS | Indicates assignment of specified processed ICD-10 code |
| DX: PHARYNGITIS | Indicates assignment of specified processed ICD-10 code |
| DX: INFLUENZA | Indicates assignment of specified processed ICD-10 code |
| DX: PNEUMONIA | Indicates assignment of specified processed ICD-10 code |
| DX: OCCLUSION AND STENOSIS | Indicates assignment of specified processed ICD-10 code |
| DX: ASTHMA | Indicates assignment of specified processed ICD-10 code |
| DX: PNEUMONITIS | Indicates assignment of specified processed ICD-10 code |
| DX: PNEUMOTHORAX | Indicates assignment of specified processed ICD-10 code |
| DX: PLEURAL EFFUSION | Indicates assignment of specified processed ICD-10 code |
| DX: RESPIRATORY FAILURE | Indicates assignment of specified processed ICD-10 code |
| DX: ULCER | Indicates assignment of specified processed ICD-10 code |
| DX: APPENDICITIS | Indicates assignment of specified processed ICD-10 code |
| DX: HERNIA | Indicates assignment of specified processed ICD-10 code |
| DX: PAIN | Indicates assignment of specified processed ICD-10 code |
| DX: HYDRONEPHROSIS | Indicates assignment of specified processed ICD-10 code |
| DX: JAUNDICE | Indicates assignment of specified processed ICD-10 code |
| DX: SYSTEMIC INFLAMMATORY RESPONSE SYNDROME | Indicates assignment of specified processed ICD-10 code |
| DX: PHLEBITIS | Indicates assignment of specified processed ICD-10 code |
| DX: SECONDARY MALIGNANT NEOPLASM | Indicates assignment of specified processed ICD-10 code |
| DX: MALIGNANT NEOPLASM | Indicates assignment of specified processed ICD-10 code |
| DX: LYMPHOMA | Indicates assignment of specified processed ICD-10 code |
| DX: LEUKAEMIA | Indicates assignment of specified processed ICD-10 code |
| DX: ENTERITIS | Indicates assignment of specified processed ICD-10 code |
| DX: PREGNANCY | Indicates assignment of specified processed ICD-10 code |
| DX: DEMENTIA | Indicates assignment of specified processed ICD-10 code |
| DX: SCHIZOPHRENIA | Indicates assignment of specified processed ICD-10 code |
| DX: DEPRESSIVE DISORDER | Indicates assignment of specified processed ICD-10 code |
| DX: SEIZURE | Indicates assignment of specified processed ICD-10 code |
| DX: HYDROCEPHALUS | Indicates assignment of specified processed ICD-10 code |
| DX: PULMONARY EMBOLISM | Indicates assignment of specified processed ICD-10 code |
| DX: ATRIAL FIBRILLATION | Indicates assignment of specified processed ICD-10 code |
| DX: HAEMORRHAGE | Indicates assignment of specified processed ICD-10 code |
| DX: LIVER DISEASE | Indicates assignment of specified processed ICD-10 code |
| DX: HEPATIC FAILURE | Indicates assignment of specified processed ICD-10 code |
| DX: PANCREATITIS | Indicates assignment of specified processed ICD-10 code |
| DX: DERMATITIS | Indicates assignment of specified processed ICD-10 code |
| DX: ARTHROPATHY | Indicates assignment of specified processed ICD-10 code |
| DX: GOUT | Indicates assignment of specified processed ICD-10 code |
| DX: LUPUS | Indicates assignment of specified processed ICD-10 code |
| DX: SCLEROSIS | Indicates assignment of specified processed ICD-10 code |
| DX: OSTEOMYELITIS | Indicates assignment of specified processed ICD-10 code |
| DX: DISCITIS | Indicates assignment of specified processed ICD-10 code |
| DX: SPONDYLOPATHIES | Indicates assignment of specified processed ICD-10 code |
| DX: MUSCLE ISCHAEMIA | Indicates assignment of specified processed ICD-10 code |
| DX: MYOSITIS | Indicates assignment of specified processed ICD-10 code |
| DX: SYNOVITIS | Indicates assignment of specified processed ICD-10 code |
| DX: BURSITIS | Indicates assignment of specified processed ICD-10 code |
| DX: FASCIITIS | Indicates assignment of specified processed ICD-10 code |
| DX: FRACTURE | Indicates assignment of specified processed ICD-10 code |
| DX: NEPHRITIC SYNDROME | Indicates assignment of specified processed ICD-10 code |
| DX: TUBULO-INTERSTITIAL DISORDERS | Indicates assignment of specified processed ICD-10 code |
| DX: NEPHRITIS | Indicates assignment of specified processed ICD-10 code |
| DX: RENAL FAILURE | Indicates assignment of specified processed ICD-10 code |
| DX: SHOCK | Indicates assignment of specified processed ICD-10 code |
| DX: OPEN WOUND | Indicates assignment of specified processed ICD-10 code |
| DX: BURN | Indicates assignment of specified processed ICD-10 code |
| DX: POISONING OR TOXIC EFFECT | Indicates assignment of specified processed ICD-10 code |
| DX: OEDEMA | Indicates assignment of specified processed ICD-10 code |
| DX: TACHYCARDIA | Indicates assignment of specified processed ICD-10 code |
| DX: BRADYCARDIA | Indicates assignment of specified processed ICD-10 code |
| DX: DELIRIUM | Indicates assignment of specified processed ICD-10 code |
| DX: OBESITY | Indicates assignment of specified processed ICD-10 code |
| DX: FEVER | Indicates assignment of specified processed ICD-10 code |
| DX: HEMATURIA | Indicates assignment of specified processed ICD-10 code |
| DX: MENTAL AND BEHAVIOURAL DISORDERS DUE TO PSYCHOACTIVE SUBSTANCE | Indicates assignment of specified processed ICD-10 code |
| DX: HOMELESSNESS OR INADEQUATE HOUSING | Indicates assignment of specified processed ICD-10 code |
| DX: OTHER CANCER | Indicates assignment of specified processed ICD-10 code |
| ORGANISM_CAND | Indicates infection by *Candida* sp. |
| ORGANISM_CNS | Indicates infection by coagulase-negative *Staphylococcus* |
| ORGANISM_EC | Indicates infection by *Escherichia coli* |
| ORGANISM_FAEM | Indicates infection by *Enterococcus faecium* |
| ORGANISM_FAES | Indicates infection by *Enterococcus faecalis* |
| ORGANISM_GAS | Indicates infection by Group A *Streptococcus* |
| ORGANISM_GBS | Indicates infection by Group B *Streptococcus* |
| ORGANISM_GCS | Indicates infection by Group C *Streptococcus* |
| ORGANISM_GDS | Indicates infection by Group D *Streptococcus* |
| ORGANISM_GGS | Indicates infection by Group G *Streptococcus* |
| ORGANISM_KO | Indicates infection by *Klebsiella oxytoca* |
| ORGANISM_KP | Indicates infection by *Klebsiella pneumoniae* |
| ORGANISM_Other Staph | Indicates infection by other *Staphylococcus* sp. |
| ORGANISM_PA | Indicates infection by *Pseudomonas aeruginosa* |
| ORGANISM_SA | Indicates infection by *Staphylococcus aureus* |
| ORGANISM_SP | Indicates infection by *Streptococcus pneumoniae* |

Table S2. Model details and performance summaries for the final models developed as part of the patient trait machine learning analysis. Model pipeline takes the format of imputation method, scaling method, feature selection method (for the full-featured models only), and classification method, with hyperparameters for each indicated in parentheses where applicable. Abbreviations: AUC, area under the curve; CCI, Charlson comorbidity index; SOFA, sequential organ failure assessment; WBC, white blood cell count; PBS, Pitt bacteremia score; RFE, recursive feature elimination.

| **Analysis** | **Model Pipeline** | **Features (count)** | **AUC [95% CI]** | **Balanced Accuracy [95% CI]** | **Accuracy [95% CI]** | **F1 Score [95% CI]** | **Sensitivity [95% CI]** | **Specificity [95% CI]** | **Precision [95% CI]** |
| --- | --- | --- | --- | --- | --- | --- | --- | --- | --- |
| **Full-Featured Models** |  |  |  |  |  |  |  |  |  |
| Mortality | Median, Min-Max, ReliefF (neighbors=10), XGBoost (max depth=2, learning rate=0.3, estimators=100) | CCI, age, SOFA, hematocrit, platelets, WBC, urea, nephritis, prothrombin, creatinine, PBS, diabetes, pneumonia, ... (99) | 0.826 [0.824–0.828] | 0.750 [0.748–0.752] | 0.769 [0.767–0.770 | 0.438 [0.436–0.441] | 0.724 [0.721–0.728] | 0.775 [0.774–0.777] | 0.314 [0.312–0.317] |
| Hospital Stay Duration | Median, Min-Max, RFE (estimator=logistic reg., step=1), XGBoost (max depth=4, learning rate=0.2, estimators=100) | sex, CCI, PBS, SOFA, dehydration, heart failure, hypokalemia, ascites, hypoxemia, endocarditis, ... (100) | 0.818 [0.817–0.819] | 0.733 [0.732–0.734] | 0.748 [0.747–0.749] | 0.664 [0.663–0.666] | 0.676 [0.674–0.678] | 0.790 [0.789–0.791] | 0.652 [0.651–0.654] |
| Healthcare Cost | Median, Min-Max, RFE (estimator=logistic reg., step=5), XGBoost (max depth=3, learning rate=0.3, estimators=100) | sex, CCI, PBS, SOFA, dehydration, heart failure, ascites, hypoxemia, endocarditis, cachexia, hypovolemia, ... (76) | 0.827 [0.825–0.828] | 0.727 [0.725–0.728] | 0.734 [0.733–0.735] | 0.689 [0.687-0.690] | 0.648 [0.646–0.650] | 0.806 [0.804–0.807] | 0.736 [0.734–0.737] |
| **Simplified Models** |  |  |  |  |  |  |  |  |  |
| Mortality | Median, Min-Max, XGBoost (max depth=2, learning rate=0.2, estimators=100) | age, CCI, PBS, SOFA (4) | 0.755 [0.753–0.758] | 0.685 [0.682–0.687] | 0.711 [0.710–0.713] | 0.359 [0.357–0.362] | 0.650 [0.645–0.654 | 0.720 [0.718–0.722] | 0.248 [0.246–0.250] |
| Hospital Stay Duration | Median, Min-Max, Random Forest (max depth=5, min samples per leaf=5, estimators=100) | sex, CCI, PBS (3) | 0.739 [0.738–0.740] | 0.671 [0.670–0.671] | 0.678 [0.677–0.679] | 0.595 [0.593–0.596] | 0.641 [0.639–0.643] | 0.700 [0.699–0.701] | 0.555 [0.553–0.556] |
| Healthcare Cost | Median, Min-Max, Random Forest (max depth=6, min samples at a leaf=20, estimators=100) | sex, CCI, PBS, SOFA (4) | 0.780 [0.779–0.782] | 0.705 [0.704–0.707] | 0.708 [0.707–0.710] | 0.678 [0.676–0.680] | 0.676 [0.673–0.678] | 0.735 [0.733–0.737] | 0.680 [0.678–0.682] |

Table S3. Model details and performance for the final machine learning models developed as part of the microbe trait-inclusive analyses in this study. The feature set column indicates the features available to the pipeline for selection: patient-related features (patient), microbe-related features (microbe), or both (integrated). Model pipeline takes the format of imputation method, scaling method, feature selection method (where applicable), and classification method, with hyperparameters for each indicated in parentheses where applicable. Unnamed genes are not included in the Features column. Abbreviations: AUC, area under the curve; CCI, Charlson comorbidity index; PBS, Pitt bacteremia score; SOFA, sequential organ failure assessment; CURB-65, confusion, urea, respiration, blood pressure, and age composite score; WBC, white blood cell count; RFE, recursive feature elimination.

| **Analysis** | **Feature Set** | **Model Pipeline** | **Features (count)** | **AUC [95% CI]** | **Balanced Accuracy [95% CI]** | **Accuracy [95% CI]** | **F1 Score [95% CI]** | **Sensitivity [95% CI]** | **Specificity [95% CI]** | **Precision [95% CI]** |
| --- | --- | --- | --- | --- | --- | --- | --- | --- | --- | --- |
| **Full Dataset Analysis** |  |  |  |  |  |  |  |  |  |  |
| *E. coli* | Patient | Median, Min-Max, ANOVA F Score, Random Forest (max depth=7, min samples per leaf=10, estimators=100) | age, sex, CCI, PBS, SOFA, CURB-65, heart failure, acidosis, prostate inflammation, ...  (99) | 0.808 [0.800–0.816] | 0.694 [0.683–0.706] | 0.775 [0.770–0.780] | 0.280 [0.268–0.292] | 0.600 [0.578–0.622] | 0.789 [0.784–0.793] | 0.183 [0.174–0.192] |
|  | Microbe | Median, Min-Max, ANOVA F Score, XGBoost (max depth=3, learning rate=0.2, estimators=10) | AmpC2, UshA, *papD*, *papC3*, *papH*, *prsF*, *papK*, *papB*, *papG*, *pdeL2*, ... (27) | 0.636 [0.625–0.647] | 0.597 [0.588–0.607] | 0.526 [0.521–0.531] | 0.173 [0.167–0.180] | 0.680 [0.661–0.699] | 0.514 [0.509–0.519] | 0.099 [0.095–0.103] |
|  | Integrated | Median, Min-Max, ANOVA F Score, XGBoost (max depth=2, learning rate=0.1, estimators=100) | age, CCI, PBS, SOFA, CURB-65, acidosis, respiratory failure, ... (26) | 0.803 [0.794–0.811] | 0.697 [0.686–0.709] | 0.781 [0.776–0.785] | 0.286 [0.274–0.297] | 0.600 [0.578–0.622] | 0.795 [0.790–0.799] | 0.188 [0.179–0.196] |
| *S. aureus* | Patient | Median, Min-Max, ANOVA F Score, Logistic Regression (C=1) | age, CCI, PBS, SOFA, CURB65, heart failure, hypoosmolality, hyperkalemia, ... (45) | 0.830 [0.825–0.835] | 0.751 [0.745–0.757] | 0.772 [0.768–0.775] | 0.452 [0.444–0.460] | 0.723 [0.712–0.734] | 0.779 [0.775–0.783] | 0.329 [0.321–0.336] |
|  | Microbe | Median, Min-Max, ReliefF (neighbors=10), Decision Tree (max depth=3, min samples per leaf=5) | *hin*, *cadC*, *blaR1-2*, *yknY2*, *bcrR*, *dnaC2*, *blaI*, *pre*, *blaZ* (65) | 0.562 [0.556–0.569] | 0.535 [0.528–0.541] | 0.543 [0.538–0.548] | 0.230 [0.223–0.236] | 0.523 [0.512–0.534] | 0.546 [0.541–0.551] | 0.147 [0.142–0.152] |
|  | Integrated | Median, Min-Max, ANOVA F Score, Logistic Regression (C=100) | age, CCI, PBS, SOFA, CURB-65, ascites, hepatorenal syndrome, hypoglycemia, ... (19) | 0.809 [0.804–0.815] | 0.717 [0.711–0.723] | 0.758 [0.754–0.761] | 0.415 [0.408–0.423] | 0.662 [0.650–0.673] | 0.772 [0.768–0.776] | 0.302 [0.296–0.310] |
| *E. faecium* | Patient | Median, Min-Max, ReliefF (neighbors=20), Random Forest (max depth=2, min samples per leaf=10, estimators=10) | CCI, bilirubin, sex, CURB65, PBS, SOFA, WBC, prothrombin, age, pneumonia, ... (89) | 0.744 [0.726–0.762] | 0.702 [0.685–0.718] | 0.732 [0.718–0.746] | 0.560 [0.535–0.585] | 0.636 [0.607–0.665] | 0.767 [0.751–0.783] | 0.500 [0.473–0.527] |
|  | Microbe | Median, Min-Max, Sequential Selection (estimator=decision tree), Multilayer Perceptron (hidden layers=[10,10], alpha=0.1) | ST1046, ST117, ST154, ST17, ST19, ST2043, ST2074, ST21, AlgC, ArgR2, Hin1, NanE2, ... (17) | 0.576 [0.555–0.596] | 0.544 [0.528–0.560] | 0.585 [0.570–0.601] | 0.370 [0.349–0.392] | 0.455 [0.428–0.481] | 0.633 [0.614–0.653] | 0.312 [0.290–0.335] |
|  | Integrated | Median, Min-Max, RFE (estimator=logistic reg., step=5), K-Neighbors (neighbors=40, weights=uniform) | *cutD*, *rpiR* ... (5) | 0.633 [0.613–0.653] | 0.570 [0.554–0.585] | 0.707 [0.695–0.720] | 0.333 [0.304–0.363] | 0.273 [0.246–0.300] | 0.867 [0.855–0.878] | 0.427 [0.387–0.470] |
| *K. pneumonia* | Patient | Median, Min-Max, ANOVA F Score, Random Forest (max depth=3, min samples per leaf=1, estimators=10) | age, CCI, PBS, acidosis, peritonitis, ascites, anoxic brain damage, myopathy, ... (19) | 0.835 [0.827–0.843] | 0.721 [0.711–0.731] | 0.783 [0.776–0.789] | 0.469 [0.456–0.482] | 0.633 [0.615–0.651] | 0.810 [0.803–0.816] | 0.372 [0.360–0.385] |
|  | Microbe | Median, Min-Max, RFE (estimator=decision tree, step=5), Multilayer Perceptron (hidden layers=[10,10], alpha=0.1) | ArgA, ArgB, ArgD, ArgE2, ArnD, AroE1, AroG, AroH, AroK, ArsA, ArsH, ArtP, Asd2, AsnA, ... (26) | 0.509 [0.496–0.522] | 0.498 [0.488–0.509] | 0.520 [0.513–0.527] | 0.228 [0.217–0.238] | 0.467 [0.446–0.488] | 0.530 [0.522–0.538] | 0.151 [0.143–0.158] |
|  | Integrated | Median, Min-Max, ANOVA F Score, Random Forest (max depth=2, min samples per leaf=10, estimators=100) | CCI, acidosis, peritonitis, anoxic brain damage, respiratory failure, ... (20) | 0.858 [0.851–0.865] | 0.747 [0.738–0.756] | 0.803 [0.798–0.808] | 0.506 [0.493–0.520] | 0.667 [0.650–0.683] | 0.827 [0.822–0.833] | 0.408 [0.395–0.422] |
| All pathogens | Patient | Median, Min-Max, ReliefF (neighbors=10), XGBoost (max depth=2, learning rate=0.3, estimators=100) | CCI, age, SOFA, hematocrit, platelets, WBC, urea, nephritis, prothrombin, creatinine, ... (99) | 0.826 [0.824–0.828] | 0.750 [0.748–0.752] | 0.769 [0.767–0.770 | 0.438 [0.436–0.441] | 0.724 [0.721–0.728] | 0.775 [0.774–0.777] | 0.314 [0.312–0.317] |
|  | Microbe | Median, Min-Max, RFE (estimator=logistic reg., step=5), Random Forest (max depth=6, min samples per leaf=5, estimators=10) | *Candida sp.*, Coag. Neg. *Staphylococcus*, *E. coli*, *E. faecium*, *E. faecalis*, Group A *Streptococcus*, ... (14) | 0.592 [0.589–0.594] | 0.568 [0.566–0.571] | 0.555 [0.553–0.556] | 0.247 [0.245–0.249] | 0.587 [0.582–0.591] | 0.550 [0.548–0.552] | 0.156 [0.155–0.158] |
|  | Integrated | Median, Min-Max, ReliefF (neighbors=20), XGBoost (max depth=2, learning rate=0.3, estimators=100) | CCI, SOFA, hematocrit, platelets, WBC, urea, age, prothrombin, bilirubin, ... (94) | 0.826 [0.824–0.828] | 0.747 [0.744–0.749] | 0.775 [0.774–0.776] | 0.440 [0.437–0.442] | 0.709 [0.705–0.713] | 0.784 [0.783–0.786] | 0.319 [0.316–0.321] |
| **Cohort Analysis** |  |  |  |  |  |  |  |  |  |  |
| Cohort 1 (*E. coli*) | Patient | Median, Min-Max, ANOVA F Score, Logistic Regression (C=0.001) | acidosis, ascites, peritonitis, cachexia, protein-calorie malnutrition, ... (11) | 0.722 | 0.647 | 0.934 | 0.308 | 0.333 | 0.962 | 0.286 |
|  | Microbe | Median, Min-Max, ReliefF (neighbors=10), Random Forest (max depth=6, min samples per leaf=20, estimators=10) | *higB2*, *entD*, *mazF*, *mazE*, *rhlE2* ... (7) | 0.654 | 0.649 | 0.632 | 0.138 | 0.667 | 0.631 | 0.077 |
|  | Integrated | Median, Min-Max, ANOVA F Score, Random Forest (max depth=4, min samples per leaf=20, estimators=100) | acidosis, ascites, cachexia, urea, *ydhC*, *dgcM*, ... (25) | 0.615 | 0.506 | 0.816 | 0.074 | 0.167 | 0.846 | 0.048 |
| Cohort 2 (*S. aureus*) | Patient | Median, Min-Max, Mutual Info, Random Forest (max depth=3, min samples per leaf=20, estimators=10) | sex, CCI, SOFA, hypoosmolality, dehydration, asphyxia, pyothorax, urine retention, ... (94) | 0.839 | 0.702 | 0.818 | 0.267 | 0.571 | 0.833 | 0.174 |
|  | Microbe | Median, Min-Max, Mutual Info, Logistic Regression (C=0.001) | Sau3AIR, YhaM, *isdC*, *metI*, *ghrB2*, *sufT*, *odhA*, *bceA1*, *macB*, ... (21) | 0.446 | 0.441 | 0.579 | 0.073 | 0.286 | 0.596 | 0.042 |
|  | Integrated | Median, Min-Max, ANOVA F Score, Random Forest (max depth=2, min samples per leaf=20, estimators=10) | intestinal obstruction, hepatorenal syndrome, falls, hematemesis, coma, fatty liver, ... (64) | 0.739 | 0.640 | 0.826 | 0.222 | 0.429 | 0.851 | 0.150 |
| Cohort 3 (all pathogens, old) | Patient | Median, Min-Max, ReliefF (neighbors=20), Random Forest (max depth=2, min samples per leaf=1, estimators=10) | age, sex, urea, CURB-65, prothrombin, pneumonia, other cancer, hematocrit, sclerosis, ... (76) | 0.828 | 0.692 | 0.870 | 0.214 | 0.500 | 0.883 | 0.136 |
|  | Microbe | Median, Min-Max, ReliefF (neighbors=20), Decision Tree (max depth=6, min samples per leaf=1) | *E. coli*, *S. pneumoniae*, Coag. Neg. *Staphylococcus*, *S. aureus*, *K. pneumoniae*, ... (11) | 0.617 | 0.539 | 0.420 | 0.076 | 0.667 | 0.411 | 0.040 |
|  | Integrated | Median, Min-Max, ReliefF (neighbors=10), Random Forest (max depth=2, min samples per leaf=10, estimators=100) | age, sex, *S. aureus*, *E. coli*, urea, Coag. Neg. *Staphylococcus*, SOFA, CURB-65, prothrombin, ... (71) | 0.653 | 0.640 | 0.769 | 0.133 | 0.500 | 0.779 | 0.077 |
| Cohort 4 (all pathogens, young) | Patient | Median, Min-Max, RFE (estimator=Logistic reg., step=5), Random Forest (max depth=5, min samples per leaf=20, estimators=10) | age, sex, SOFA, CURB-65, dehydration, hypoosmolality, hypokalemia, ... (49) | 0.948 | 0.859 | 0.916 | 0.296 | 0.800 | 0.919 | 0.182 |
|  | Microbe | Median, Min-Max, PCA (components=4), K-Neighbors (neighbors=40, weights=distance) | All species classification features, decomposed | 0.438 | 0.387 | 0.757 | 0.000 | 0.000 | 0.774 | NaN |
|  | Integrated | Median, Min-Max, RFE (estimator=Decision Tree, step=1), Random Forest (max depth=6, min samples per leaf=10, estimators=10) | SOFA, CURB-65, dehydration, hypoosmolality, heart failure, hypokalemia, acidosis, ... (100) | 0.756 | 0.652 | 0.894 | 0.143 | 0.400 | 0.905 | 0.087 |
| **Targeted Analysis** |  |  |  |  |  |  |  |  |  |  |
| *papG*  (*E. coli*) | Patient | Median, Min-Max, XGBoost (max depth=2, learning rate=0.2, estimators=100) | age, CCI, PBS, SOFA | 0.695 | 0.620 | 0.740 | 0.212 | 0.480 | 0.760 | 0.136 |
|  | Microbe | Median, Min-Max, Logistic Regression (C=100) | *papG* | 0.591 | 0.591 | 0.447 | 0.167 | 0.760 | 0.423 | 0.094 |
|  | Integrated | Median, Min-Max, XGBoost (max depth=2, learning rate=0.1, estimators=100) | age, CCI, PBS, SOFA, *papG* | 0.710 | 0.611 | 0.757 | 0.210 | 0.440 | 0.782 | 0.138 |
| KdpC_1  (*S. aureus*) | Patient | Median, Min-Max, XGBoost (max depth=2, learning rate=0.1, estimators=100) | age, CCI, PBS, SOFA | 0.781 | 0.714 | 0.741 | 0.406 | 0.677 | 0.751 | 0.290 |
|  | Microbe | Median, Min-Max, Random Forest (max depth=2, min samples per leaf=10, estimators=100) | KdpC_1 | 0.535 | 0.513 | 0.665 | 0.193 | 0.308 | 0.719 | 0.141 |
|  | Integrated | Median, Min-Max, XGBoost (max depth=2, learning rate=0.1, estimators=100) | age, CCI, PBS, SOFA, KdpC_1 | 0.788 | 0.694 | 0.717 | 0.379 | 0.662 | 0.726 | 0.266 |
| VanX  (*E. faecium*) | Patient | Median, Min-Max, Logistic Regression (C=100) | age, CCI, PBS, SOFA | 0.733 | 0.627 | 0.707 | 0.455 | 0.455 | 0.800 | 0.455 |
|  | Microbe | Median, Min-Max, Logistic Regression (C=100) | VanX | 0.483 | 0.412 | 0.561 | 0.100 | 0.091 | 0.733 | 0.111 |
|  | Integrated | Median, Min-Max, Logistic Regression (C=1) | age, CCI, PBS, SOFA, VanX | 0.715 | 0.520 | 0.634 | 0.286 | 0.273 | 0.767 | 0.300 |
| *K. pneumonia* virulence score | Patient | Median, Min-Max, Logistic Regression (C=100) | age, CCI, PBS, SOFA | 0.763 | 0.714 | 0.677 | 0.418 | 0.767 | 0.661 | 0.287 |
|  | Microbe | Median, Min-Max, Logistic Regression (C=100) | virulence score | 0.438 | 0.448 | 0.621 | 0.138 | 0.200 | 0.696 | 0.105 |
|  | Integrated | Median, Min-Max, Random Forest (max depth=2, min samples per leaf=5, estimators=10) | age, CCI, PBS, SOFA, virulence score | 0.735 | 0.692 | 0.803 | 0.451 | 0.533 | 0.851 | 0.391 |
| All *S. aureus* virulence proteins | Patient | Median, Min-Max, Mutual Info, XGBoost (max=2, learning rate=0.1, estimators=100) | age, CCI, PBS, SOFA ... (4) | 0.781 | 0.714 | 0.741 | 0.406 | 0.677 | 0.751 | 0.290 |
|  | Microbe | Median, Min-Max, ANOVA F Score, Random Forest (max depth=2, min samples per leaf=1, estimators=10) | SdrD, EntH, ... (4) | 0.565 | 0.520 | 0.563 | 0.216 | 0.462 | 0.578 | 0.141 |
|  | Integrated | Median, Min-Max, ANOVA F Score, Random Forest (max depth=4, min samples per leaf=10, estimators=100) | age, CCI, PBS, SOFA, ... (5) | 0.783 | 0.723 | 0.689 | 0.392 | 0.769 | 0.677 | 0.263 |

Table S4. Summary metrics for the *Calgary BSI Cohort* datasets used for the analyses in this study. The *E. coli, S. aureus, E. faecium,* and *K. pneumoniae* datasets were used to assess microbial multi-omic traits as predictors of BSI clinical outcome, while the All-Pathogens dataset was used for the patient trait and species classification analyses.

| **Dataset** | **# Samples Pre-Processing** | **# Samples Post-Processing** | **# Patient Features** | **# Microbial Genomic Features** | **# Microbial Proteomic Features** | **Age (Mean ± SD) (years)** | **Sex (% Female)** | **CCI Mean ± SD)** | **30-Day Mortality Incidence (%)** |
| --- | --- | --- | --- | --- | --- | --- | --- | --- | --- |
| *E. coli* | 10,501 | 6,029 | 184 | 68,192 | 2,582 | 54.6 ± 15.8 | 55.4 | 2.6 ± 3.0 | 8.9 |
| *S. aureus* | 8,078 | 4,737 | 256 | 8,863 | 1,805 | 50.3 ± 17.9 | 34.6 | 2.6 ± 2.8 | 12.3 |
| *E. faecium* | 844 | 552 | 316 | 11,380 | 1,958 | 52.4 ± 18.3 | 38.0 | 3.1 ± 2.7 | 27.5 |
| *K. pneumoniae* | 2,427 | 1,555 | 251 | 55,942 | 2,672 | 54.5 ± 16.1 | 44.2 | 3.2 ± 3.0 | 14.3 |
| All-Pathogens | 35,746 | 20,930 | 242 | N/A | N/A | 52.0 ± 17.6 | 43.6 | 2.6 ± 2.9 | 14.9 |

Table S5. Assembly quality metrics. SA = *Staphylococcus aureus*. FAEM = *Enterococcus faecium*, EC = *Escherichia coli*, KP = *Klebsiella pneumoniae*.

| **Metric** | **SA** | **FAEM** | **EC** | **KP** |
| --- | --- | --- | --- | --- |
| **Genome Size (+/- 20%)** | 2730000 | 2840000 | 5000000 | 5390000 |
| **GC Content (+/- 10%)** | 36.0 | 40.6 | 52.4 | 58.5 |
| **Annotated genes (+/- 20%)** | 2530 | 2740 | 4700 | 5000 |
| **Maximum assembly contigs** | 200 | 300 | 300 | 300 |
| **Minimum assembly contig N50** | 25000 | 15000 | 25000 | 25000 |
| **Minimum sequencing coverage** | 20 | 20 | 20 | 20 |
| **Minimum trimmed read length** | 120 | 120 | 120 | 120 |
| **Minimum mean base quality** | 30 | 30 | 30 | 30 |
| **Max StrainGST strains (score>0.2)** | 1 | 1 | 1 | 1 |
